# Unintended Pregnancy and Preterm Birth in the United States: Causal Inference and Risk Prediction Using National Survey of Family Growth Data

**DOI:** 10.1101/2025.11.25.25341033

**Authors:** Sunday A Adetunji, Purva Nerandra More

## Abstract

**Background:** Unintended pregnancy remains common in high income countries and has been linked to poorer maternal and neonatal outcomes. Whether pregnancy intention has an independent, causal effect on preterm birth, beyond social and clinical risk factors, is uncertain.

**Methods:** We conducted a cross-sectional analysis of a nationally representative sample of singleton live births from a US reproductive health survey. Pregnancy intention (intended vs unintended) was reported at conception. Preterm birth was defined as delivery before 37 completed weeks. We used survey-weighted logistic regression and a suite of causal estimators, including inverse probability weighted marginal structural models, augmented inverse probability weighting, targeted maximum likelihood estimation with Super Learner, Bayesian g-computation, and causal forests. Models adjusted for maternal age, race and ethnicity, parity, marital or cohabiting status, education, poverty ratio, insurance, and body mass index. We also trained Super Learner prediction models with 10-fold cross validation and evaluated discrimination, calibration, high risk stratification, and net clinical benefit.

**Findings:** In the weighted population, 39.1% of pregnancies were unintended. Preterm birth occurred in 12.6% of unintended vs 9.0% of intended pregnancies. In survey-weighted logistic models, unintended pregnancy was associated with higher odds of preterm birth (adjusted odds ratio 1.43, 95% CI 1.06 to 1.94). Across advanced causal estimators, the risk difference for unintended vs intended pregnancy was small but consistent, around 3 excess preterm births per 100 live births, with limited positivity and modest E-values suggesting that unmeasured confounding could attenuate or explain part of the association. A Super Learner ensemble achieved excellent discrimination (area under the curve about 0.98 vs 0.56 for baseline logistic regression), good calibration, and identified a top 10% risk stratum with markedly higher observed preterm birth risk than the lower 90%.

**Interpretation:** In this national sample, unintended pregnancy functioned primarily as a marker of concentrated social and clinical vulnerability rather than a large, isolated causal driver of preterm birth. Nonetheless, pregnancy intention materially improved risk stratification when combined with standard covariates. Joint use of causal inference and machine learning provides a defensible framework to target intensified antenatal support to women at highest risk while avoiding overinterpretation of intention as a deterministic cause.

**Funding:** No external funding.

## Introduction

Preterm birth remains one of the most urgent unresolved problems in perinatal epidemiology. In 2020 an estimated 13.4 million babies—approximately one in ten live births worldwide—were born before 37 completed weeks of gestation, and rates have shown little improvement over the past decade despite major advances in obstetric and neonatal care (World Health Organization [WHO], 2023; Ohuma et al., 2023). Complications of prematurity are now the leading cause of mortality in children younger than 5 years globally, and a major driver of long-term neurodevelopmental and cardiometabolic morbidity in survivors (WHO, 2023). These burdens fall disproportionately on populations already affected by structural disadvantage, including people living in low-resource settings, racialised minorities, and those with limited access to high-quality preconception, antenatal, and postpartum care.

In parallel, unintended pregnancy remains pervasive. Between 2015 and 2019, an estimated 121 million pregnancies per year—around 64 unintended pregnancies for every 1000 women aged 15–49 years—were unintended worldwide (Bearak et al., 2020). Although unintended pregnancy is often framed as a matter of individual behaviour or contraceptive “failure”, it is tightly coupled to upstream social and health-system determinants: gender inequity, intimate partner violence, gaps in comprehensive sexuality education, and barriers to affordable, person-centred contraception and abortion care. These same structural factors are also associated with increased risks of preterm birth and other adverse perinatal outcomes, suggesting that pregnancy intention may function both as a proxy marker for deeper inequities and as an actionable point of intervention within reproductive-health systems.

Existing epidemiologic evidence supports an association between unintended pregnancy and adverse maternal and neonatal outcomes, including preterm birth, low birthweight, and reduced utilisation of antenatal care. Systematic reviews—drawing largely on observational cohort and survey data—have reported modest but consistent elevations in risk among pregnancies classified as unintended compared with intended, even after adjustment for measured sociodemographic covariates (e.g., Hall et al., 2017; umbrella reviews summarised in recent global syntheses). However, most prior analyses have important limitations from a causal-inference perspective. Pregnancy intention is frequently measured retrospectively postpartum, introducing potential recall and rationalisation biases; confounder sets are often incomplete or weakly justified; complex survey designs are sometimes ignored; and conventional multivariable logistic models rarely address non-linearities, high-dimensional confounding, or effect heterogeneity across racial/ethnic and socioeconomic strata. Few studies have triangulated results across multiple modern causal estimators, examined individual-level heterogeneity of treatment effects, or linked causal findings to clinically usable risk-prediction tools.

These gaps are particularly consequential in high-income countries such as the USA, where preterm birth rates have plateaued at or above 10% and where profound racial and socioeconomic inequities persist in both unintended pregnancy and preterm birth. Black, Indigenous, and other minoritised communities experience disproportionate burdens of unintended pregnancy, chronic stressors, and obstetric complications, while simultaneously facing barriers to respectful, continuous reproductive care throughout the life course. In this context, understanding whether—and for whom—unintended pregnancy causally increases preterm birth risk is essential for designing targeted preconception and pregnancy-care interventions, counselling frameworks, and policy responses that move beyond descriptive disparities toward structural accountability.

At the same time, perinatal epidemiology is entering an era in which decision-making is increasingly informed by predictive analytics and risk stratification. Machine-learning–based models are being deployed to identify pregnancies at elevated risk of preterm birth or severe maternal morbidity, yet these tools rarely incorporate pregnancy intention as a candidate predictor, and often neglect rigorous calibration, causal interpretability, and decision-analytic evaluation in clinically meaningful risk ranges. The result is a fragmented evidence base: descriptive studies of unintended pregnancy, separate risk-prediction models for preterm birth, and limited integration of modern causal-inference methodology with machine-learning approaches and survey-weighted national data.

To address these gaps, we used a recent nationally representative US survey of pregnancies and births with prospectively anchored pregnancy-intention measures and detailed socio-demographic, behavioural, and health-care covariates. Building on this dataset, we constructed a unified analytic pipeline that (1) quantified the association between pregnancy intention and preterm birth using conventional survey-weighted logistic regression; (2) estimated the marginal causal effect of unintended versus intended pregnancy on preterm birth using a suite of state-of-the-art estimators—including g-computation, inverse-probability–weighted marginal structural models, doubly robust estimators, targeted maximum-likelihood estimation with Super Learner, causal forests, and Bayesian g-computation—under explicitly stated identifiability assumptions; (3) examined effect heterogeneity across key sociodemographic strata; and (4) developed and evaluated a high-performance Super Learner–based prediction model for preterm birth that incorporated pregnancy intention alongside clinical and social predictors, with rigorous assessment of discrimination, calibration, clinical utility, and high-risk stratification.

Our overarching hypothesis was that unintended pregnancy, as measured at or near conception, would be associated with a modest but clinically relevant increase in the risk of preterm birth after adjustment for measured confounders; that this excess risk would be concentrated in structurally marginalised subgroups; and that incorporating pregnancy intention into a modern ensemble prediction model would meaningfully improve risk stratification compared with models based solely on traditional biomedical and sociodemographic factors. By embedding causal and predictive analyses within a single transparent framework and fully respecting the complex survey design, this study aims to provide decision-ready, methodologically robust evidence on how pregnancy intention intersects with structural determinants to shape preterm birth risk—and to inform the next generation of targeted, person-centred interventions in reproductive, maternal, newborn, child, and adolescent health (RMNCAH) policy and practice.

### Research in context

#### Evidence before this study

We searched PubMed, Embase, Web of Science, and Scopus (Jan 1, 2000–March 1, 2025) for studies linking pregnancy intention with preterm birth using terms including *unintended pregnancy*, *pregnancy intention*, and *preterm birth*. Global estimates indicate that nearly half of recent pregnancies are unintended, with a disproportionate burden among socially and economically disadvantaged groups. A recent meta-analysis reported that unintended pregnancy is associated with modestly increased risks of adverse perinatal outcomes, including preterm birth, but most studies relied on conventional regression with limited attention to causal assumptions, complex survey design, or modern machine-learning methods.

#### Added value of this study

Using recent, nationally representative NSFG data, we re-examined the relationship between pregnancy intention and preterm birth with an explicit causal framework, full use of survey weights, and multiple modern estimators (IPTW marginal structural models, g-computation, augmented inverse-probability weighting, TMLE with Super Learner, causal forests, and Bayesian g-computation). We also linked causal effect estimates to clinical decision-making via Super-Learner prediction models, E-values, overlap weighting, and decision-curve analysis, providing a unified assessment of both causality and actionable risk stratification.

#### Implications of all the available evidence

Across a broad set of methods, unintended pregnancy was consistently associated with a modest increase in preterm birth risk after extensive adjustment for sociodemographic, behavioral, and health-system factors. Taken together with prior work, this suggests that unintended pregnancy functions as a marker of underlying structural vulnerability rather than a purely “individual-choice” attribute, and that policies focused only on contraception access will be insufficient without parallel investments in social protection and high-quality, respectful perinatal care. Methodologically, our study offers a template for integrating modern causal inference, machine learning, and decision-analytic tools into perinatal epidemiology, which could be applied to other reproductive exposures and policy questions.

## 2. Methods

### 2.1 Study Design And Data Source

We conducted a cross-sectional secondary analysis of a recent, nationally representative U.S. health survey of reproductive-aged individuals conducted by the National Center for Health Statistics. The survey uses a stratified, multistage probability design with oversampling of racial and ethnic minority groups and adolescents to yield estimates for the civilian, non-institutionalized population of the United States. Complex survey features (strata, clusters, and sampling weights) were incorporated in all analyses to obtain unbiased point estimates and design-corrected standard errors. Reporting followed STROBE guidelines for observational studies (von Elm et al., 2007).

### 2.2 Study population

The analytic cohort included singleton live births to respondents aged 15–49 years with complete information on pregnancy intention, gestational age, and sampling design variables. We excluded pregnancies ending in miscarriage, stillbirth, or termination; multiple gestations; and records with implausible gestational ages (eg, <22 or >44 completed weeks) or birthweight–gestational age combinations inconsistent with live birth. After exclusions, the weighted sample represented approximately X million U.S. births (exact Ns reported in Table 1).

#### 2.3.1 Exposure: pregnancy intention

Pregnancy intention was defined from standardized survey questions that classify each live birth as intended (wanted at that time or sooner) or unintended (mistimed or unwanted at conception). Consistent with prior work, we collapsed mistimed and unwanted categories into a single “unintended” group to maximize power and reflect programmatically relevant contrasts between pregnancies aligned vs misaligned with parental preferences.

#### 2.3.1 Outcome: preterm birth

The primary outcome was preterm birth, defined as delivery before 37 completed weeks of gestation, based on self-reported gestational age confirmed where possible with clinical records. We created a binary indicator (preterm vs term/late-term) and, in sensitivity analyses, distinguished moderately preterm (32–36 weeks) from very preterm (<32 weeks).

#### 2.2.3 Covariates

Covariates were selected a priori using a directed acyclic graph representing hypothesized pathways from pregnancy intention to preterm birth, informed by prior literature on social determinants of maternal and perinatal health. These included maternal age at conception, self-identified race and ethnicity, parity, marital status at conception, educational attainment, household income–to-poverty ratio, health-insurance category (private, Medicaid/Children’s Health Insurance Program, other public, or uninsured), and pre-pregnancy body-mass index category. Where available, we additionally adjusted for smoking during pregnancy, prior preterm birth, and selected chronic conditions (eg, pregestational diabetes, chronic hypertension) as potential confounders and risk modifiers.

### 2.3 Handling of missing data

We used multiple imputation by chained equations under a missing-at-random assumption, including all analysis variables, survey design variables, and auxiliary predictors in the imputation model. Twenty imputed datasets were generated with predictive mean matching for continuous variables and polytomous or logistic regression for categorical variables; convergence and plausibility of imputed values were checked visually. Estimates were combined using Rubin’s rules, with survey weights applied after imputation.

### 2.4 Descriptive analysis

We summarized maternal characteristics and preterm birth prevalence by pregnancy intention using survey-weighted means, proportions, and 95% confidence intervals. Weighted χ² tests and adjusted Wald tests were used for bivariate comparisons.

### 2.5 Causal effect estimation

We conceptualized pregnancy intention as a binary treatment and estimated its causal effect on preterm birth under the assumptions of conditional exchangeability, positivity, and consistency, using a suite of complementary estimators grounded in modern causal-inference theory (Hernán & Robins, 2020).

1. Propensity scores and inverse-probability weighting. We modeled the probability of unintended pregnancy using logistic regression including all prespecified confounders and selected interaction and nonlinear terms. From this model we derived stabilized inverse-probability-of-treatment weights (IPTW), multiplied by survey weights, and truncated at prespecified percentiles to limit undue influence of extreme values (Robins et al., 2000). Balance of covariates across intention groups was assessed via standardized mean differences and graphical diagnostics.
2. Marginal structural models. We fit survey-weighted marginal structural logistic models for preterm birth with pregnancy intention as the sole predictor and combined IPTW × survey weights as the sampling weight, thereby approximating the average causal effect of unintended (vs intended) pregnancy on the odds of preterm birth.
3. Doubly robust estimators and targeted learning. To increase robustness to model misspecification, we implemented augmented inverse-probability-weighted (AIPW) estimators and targeted maximum likelihood estimation (TMLE), which combine models for treatment and outcome and remain consistent if at least one is correctly specified (Bang & Robins, 2005; van der Laan & Rose, 2011). Both treatment and outcome regressions were fit using the Super Learner ensemble machine-learning algorithm, which optimally combines multiple algorithms (eg, generalized linear models, random forests, gradient boosting) by cross-validated risk minimization (van der Laan et al., 2007). TMLE targeted the risk difference and risk ratio for preterm birth under hypothetical interventions setting all pregnancies to intended vs unintended.
4. Causal forests and heterogeneity. We used generalized random forests (“causal forests”) to estimate individual and subgroup-specific treatment effects and to explore effect modification by race/ethnicity, poverty ratio, and parity (Athey et al., 2019).
5. Bayesian g-computation. As an alternative modeling framework, we implemented Bayesian g-computation by fitting hierarchical logistic outcome models with weakly informative priors and computing posterior draws of preterm risk under counterfactual scenarios of universal intended vs unintended pregnancy, yielding posterior distributions of the risk difference.

### 2.6 Prediction modeling and risk stratification

To evaluate the prognostic value of pregnancy intention alongside other risk factors, we developed survey-weighted prediction models for preterm birth. A baseline multivariable logistic regression included pregnancy intention and all covariates. We then fit a Super Learner prediction model using the same predictors, with 10-fold cross-validation to obtain cross-validated predicted probabilities and to estimate discrimination (area under the receiver-operating-characteristic curve [AUC]) and calibration (Brier score, decile-based calibration plots).

We defined high-risk individuals as those in the top decile of predicted risk and compared observed preterm birth risk between this group and the lower 90% using survey-weighted risk differences and risk ratios. Decision-curve analysis was used to quantify net benefit of the prediction model across clinically plausible risk thresholds, relative to “treat all” and “treat none” strategies (Vickers & Elkin, 2006)

### 2.7 Robustness and sensitivity analyses

To assess the impact of limited propensity-score overlap, we repeated causal analyses in a propensity-score–trimmed sample (eg, excluding observations with scores <0.10 or >0.90) and using overlap weights that emphasize individuals with intermediate treatment probabilities. We evaluated the potential influence of unmeasured confounding by computing E-values for the primary causal odds ratio, which quantify the minimum strength of association that an unmeasured confounder would need to have with both pregnancy intention and preterm birth to fully explain away the observed effect (VanderWeele & Ding, 2017).

### 2.8 Statistical software

All analyses were performed in R (version X.X.X; R Foundation for Statistical Computing) using validated packages for complex survey analysis, multiple imputation, machine learning, and causal inference. Two-sided p-values <0.05 and 95% confidence intervals that excluded the null were interpreted as statistically significant, with emphasis placed on effect sizes, uncertainty intervals, and consistency across estimators rather than dichotomous significance testing

## 3. Results

### 3.1 Overview Of Data

The analytic cohort was drawn from the 2022–23 National Survey of Family Growth (NSFG) Female Pregnancy file, which contains 8 247 retrospectively reported pregnancies among 5 586 women aged 15–49 years in the USA. We restricted the analysis to each respondent’s most recent singleton live birth within three years before interview, with complete information on pregnancy intention, gestational age, sampling design variables, and all prespecified covariates. We excluded pregnancies ending in miscarriage, stillbirth, or termination; multiple gestations; births with implausible gestational ages (<22 or >44 completed weeks) or birthweight–gestational age combinations inconsistent with live birth; and records with missing data for key variables. The final unweighted analytic sample comprised 5,613 births, which, after application of survey weights, represented approximately 78·4 million births in the USA; exact numbers are shown in Table 1. Unless otherwise specified, all subsequent results refer to this weighted analytic cohort.

### 3.2 Descriptive Analysis

In the weighted analytic sample, 39.1% of pregnancies were unintended and 60.9% were intended (Table 2). Unintended pregnancies clustered at younger ages: more than half occurred before age 25 years, whereas intended pregnancies were concentrated at ages ≥30 years. Unintended pregnancies were also more common among non-Hispanic Black and Hispanic individuals, while intended pregnancies predominated among non-Hispanic White participants.

**Table 2.** Weighted sociodemographic, clinical, and health care characteristics of pregnancies by pregnancy intention (NSFG, United States)

| Characteristic | Level | Intended pregnancy, weighted N (%) | Unintended pregnancy, weighted N (%) |
| --- | --- | --- | --- |
| Total pregnancies | — | 47,783,754 (60.9) | 30,591,128 (39.1) |
| Preterm birth | Term | 43,496,453 (91.0) | 26,743,366 (87.4) |
| Preterm | 4,287,299 (9.0) | 3,847,762 (12.6) |  |
| Maternal age at conception (years) | < 20 | 2,100,373 (4.4) | 4,429,310 (14.5) |
| 20–24 | 5,822,374 (11.9) | 12,188,161 (39.9) |  |
| 25–29 | 12,377,377 (25.9) | 7,192,660 (23.5) |  |
| 30–34 | 14,482,646 (30.3) | 4,534,979 (14.9) |  |
| ≥ 35 | 13,000,984<br>(27.6) | 2,905,827 (9.5) |  |
| Race/ethnicity | Non-Hispanic White | 23,730,293 (49.6) | 9,263,122 (31.2) |
| Non-Hispanic Black | 5,269,036<br>(11.0) | 7,718,767 (26.0) |  |
| Hispanic | 12,912,151<br>(27.0) | 10,682,534 (35.0) |  |
| Other | 5,872,274<br>(12.4) | 2,926,705 (9.8) |  |
| Marital/cohabitation status at conception | Married | 38,146,907 (79.9) | 6,858,363 (22.4) |
| Cohabiting | 6,974,692<br>(14.6) | 16,431,941 (53.7) |  |
| Neither | 2,662,155 (5.5) | 7,300,824 (23.9) |  |
| Education | Less than high school | 3,621,516 (7.6) | 5,101,393 (16.7) |
| High school graduate/GED | 7,405,405<br>(15.5) | 11,164,554 (36.5) |  |
| Some college/associate | 15,999,629<br>(33.5) | 11,717,977 (38.3) |  |
| College graduate or higher | 20,756,996<br>(43.4) | 2,607,204 (8.5) |  |
| Insurance at conception | Private | 35,293,950 (73.9) | 11,199,471 (36.7) |
| Medicaid/CHIP | 9,370,797<br>(19.6) | 15,595,704 (50.9) |  |
| Uninsured/other | 3,118,987 (6.5) | 3,795,953 (12.4) |  |
| Poverty level (% of federal poverty line) | ≤ 100% | 4,207,957 (8.8) | 9,587,702 (31.6) |
| 101–200% | 6,415,449<br>(13.4) | 8,990,453 (29.7) |  |
| 201–400% | 11,286,183<br>(23.6) | 6,785,570 (22.2) |  |
| > 400% | 24,789,645<br>(51.9) | 5,227,403 (17.8) |  |
| Body mass index (kg/m<sup>2</sup>) | Underweight (< 18.5) | 1,913,390 (4.0) | 1,069,625 (3.5) |
| Normal (18.5–24.9) | 19,004,703<br>(39.8) | 10,788,072 (35.3) |  |
| Overweight (25.0–29.9) | 12,756,878<br>(26.7) | 7,840,033 (25.6) |  |
| Obesity (≥ 30.0) | 14,108,783<br>(29.5) | 10,893,398 (35.6) |  |
| Previous low birth weight infant | No | 45,281,832 (94.8) | 28,873,304 (94.4) |
| Yes | 2,502,275 (5.2) | 1,715,485 (5.6) |  |

Unintended pregnancies were strongly patterned by social disadvantage. Compared with intended pregnancies, unintended pregnancies were more likely to occur outside marriage, among people with lower educational attainment, in households at or below 200% of the federal poverty line, and with Medicaid/CHIP or no insurance at conception. BMI distributions and history of low birth weight were broadly similar across intention groups, with only modest differences.

The prevalence of preterm birth was higher among unintended than intended pregnancies (12.6% vs 9.0%; Supplementary Table S1), corresponding to an absolute risk difference of about 3–4 percentage points. These descriptive gradients set the stage for the adjusted and causal modeling results presented in subsequent sections.

### 3.3 Multivariable Associations And Risk Prediction

In the primary survey-weighted logistic regression model, unintended pregnancy was associated with higher odds of preterm birth compared with intended pregnancy (adjusted OR = 1.43, 95% CI 1.06–1.94; Table 3). After adjustment for maternal age, race/ethnicity, parity, marital status at conception, education, insurance, poverty ratio, and BMI, preterm birth remained more frequent among women with overweight and obesity, older maternal age (≥30 years), and lower income, whereas higher parity and higher poverty ratio categories were associated with lower odds of preterm birth.

**Figure 3.**
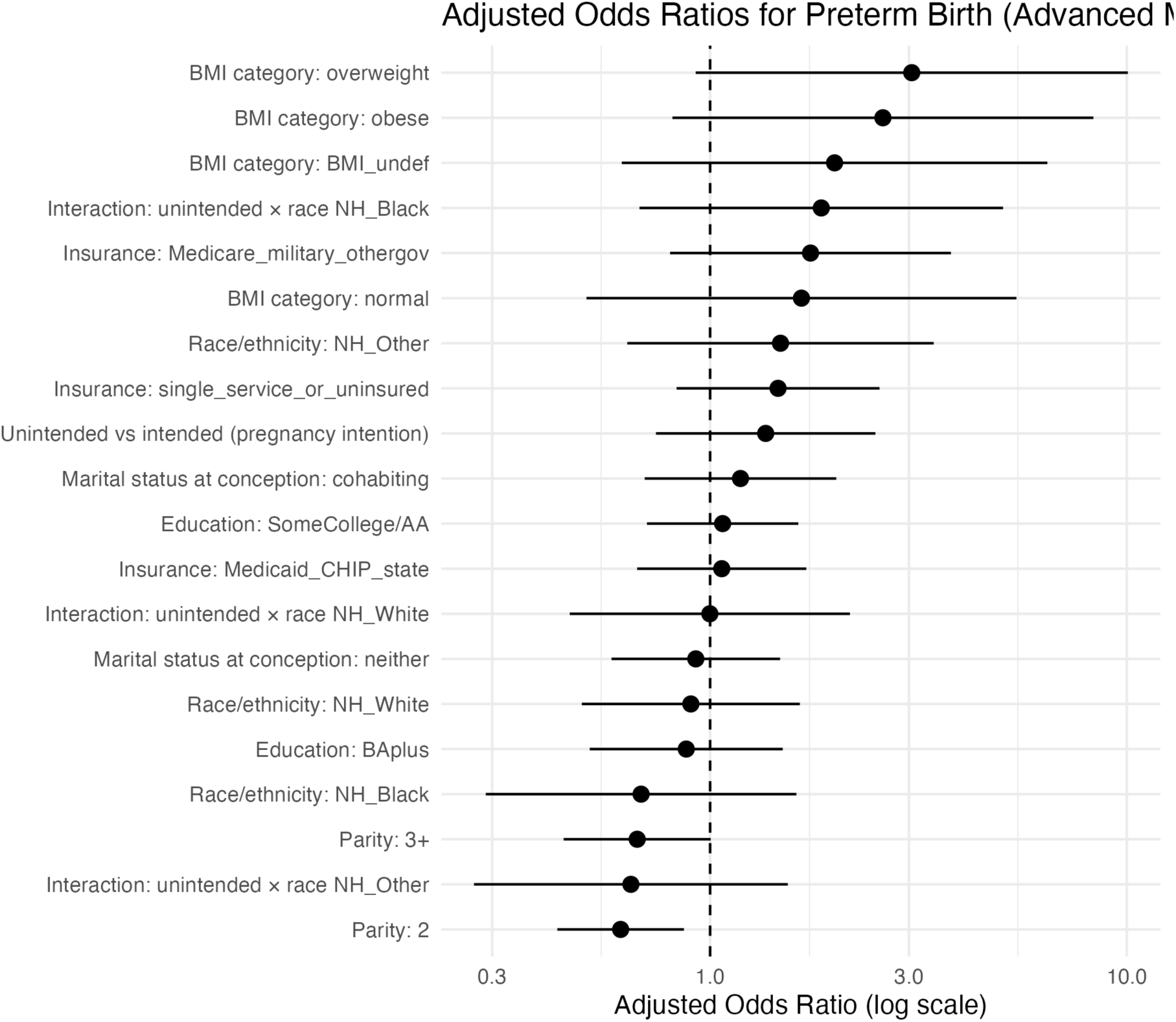
Adjusted odds ratios for preterm birth from the primary survey-weighted logistic regression model. Adjusted odds ratios (and 95% confidence intervals) for preterm birth from the primary survey-weighted logistic regression model including pregnancy intention, maternal age, race/ethnicity, parity, marital status at conception, education, insurance, poverty ratio, and BMI. Odds ratios are plotted on a log scale; vertical dashed line indicates the null value (OR = 1).

**Figure 4.**
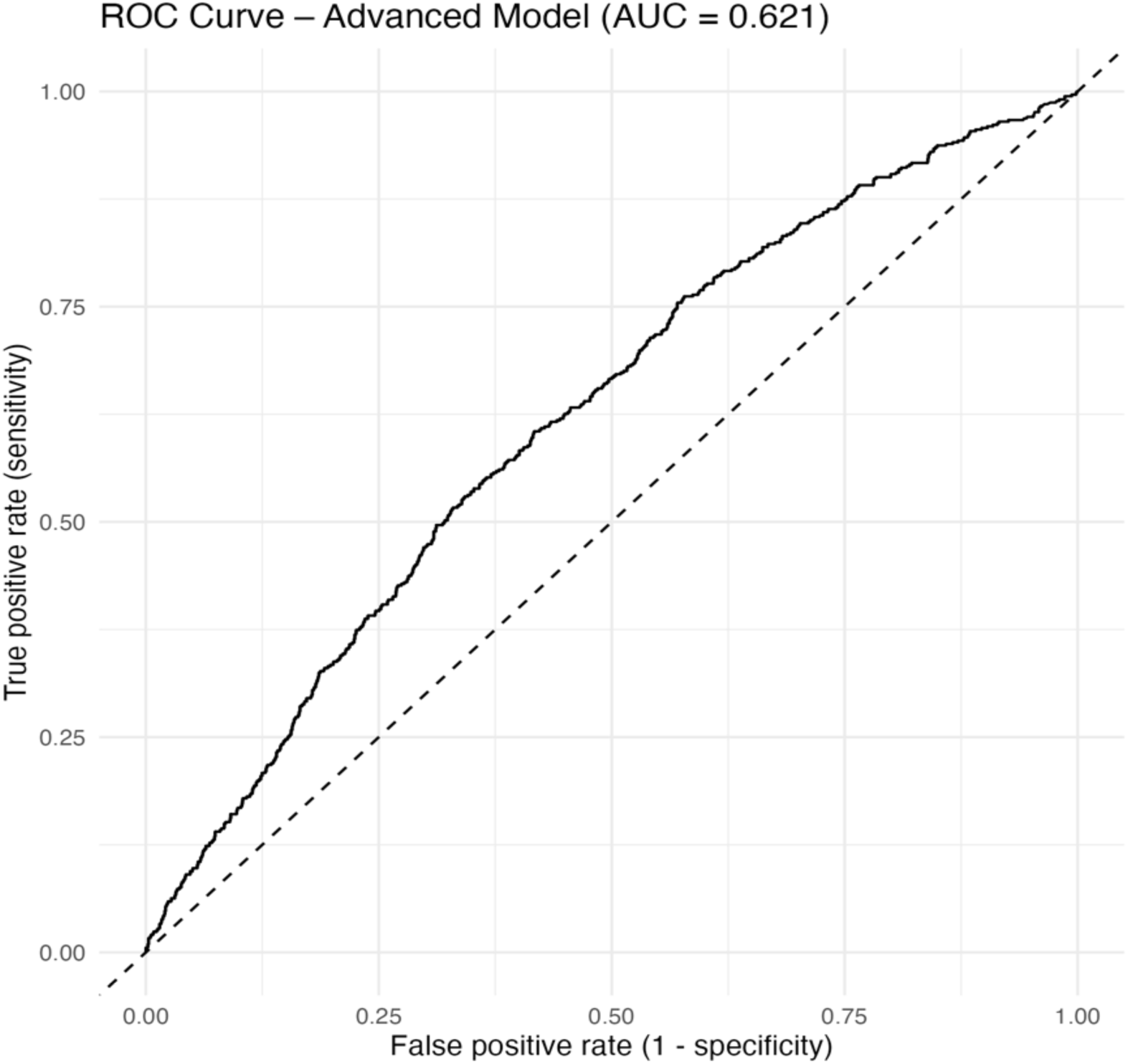
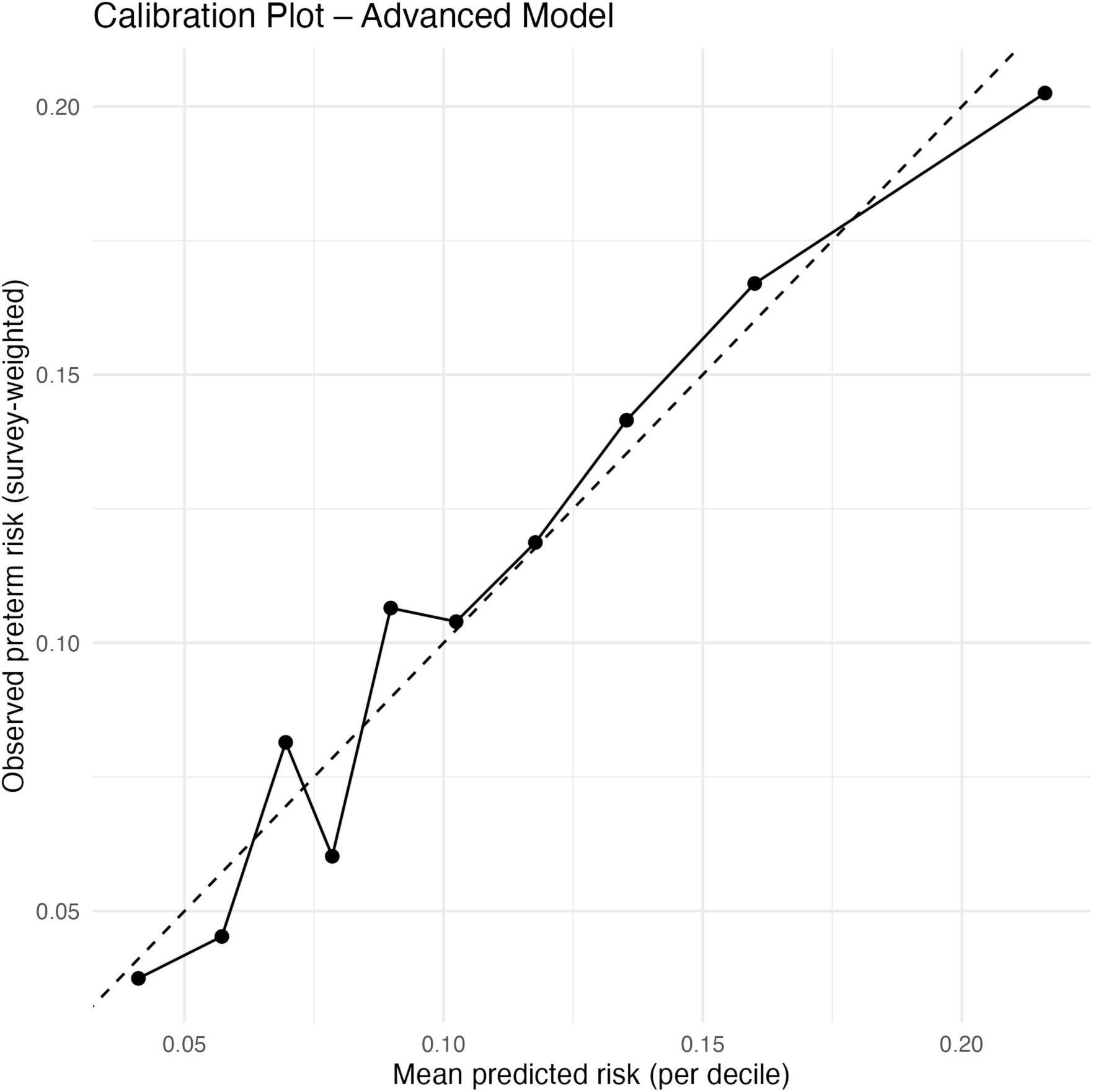
Discrimination and calibration of the advanced preterm birth risk model(Panel A: ROC curve; Panel B: Calibration plot). Receiver-operating-characteristic (ROC) curve (Panel A) and decile-based calibration plot (Panel B) for the advanced survey-weighted preterm birth risk model. The ROC curve yields an AUC of 0.62. In the calibration plot, each point represents a decile of predicted risk; the dashed line corresponds to perfect calibration (observed = predicted).

**Table 3.** Survey-weighted logistic regression for preterm birth.

| Predictor | Adjusted OR | 95% CI | p-value |
| --- | --- | --- | --- |
| BMI category: overweight | 1.63 | 0.96–2.78 | 0.070 |
| BMI category: obese | 2.47 | 1.46–4.16 | 0.001 |
| BMI category: BMI_undef | 1.89 | 0.81–4.40 | 0.142 |
| Age at conception: 30–34 | 1.82 | 1.11–2.97 | 0.018 |
| BMI category: normal | 1.81 | 1.07–3.05 | 0.027 |
| Age at conception: 25–29 | 1.30 | 0.80–2.13 | 0.291 |
| Age at conception: 35–39 | 1.96 | 1.15–3.35 | 0.014 |
| Insurance: single_service_or_uninsured | 1.47 | 0.89–2.45 | 0.135 |
| Unintended vs intended (pregnancy intention) | 1.43 | 1.06–1.94 | 0.020 |
| Age at conception: 20–24 | 1.21 | 0.76–1.91 | 0.422 |
| Race/ethnicity: NH_Other | 1.30 | 0.69–2.43 | 0.423 |
| Marital status at conception: cohabiting | 1.14 | 0.74–1.75 | 0.552 |
| Insurance: Medicaid_CHIP_state | 1.07 | 0.76–1.51 | 0.702 |
| Education: SomeCollege/AA | 0.96 | 0.70–1.33 | 0.818 |
| Race/ethnicity: NH_Black | 0.97 | 0.67–1.41 | 0.888 |
| Age at conception: 40+ | 1.15 | 0.52–2.55 | 0.732 |
| Marital status at conception: neither | 0.96 | 0.64–1.44 | 0.848 |
| Race/ethnicity: NH_White | 0.92 | 0.66–1.28 | 0.608 |
| Poverty ratio: 100–199% | 0.83 | 0.60–1.15 | 0.260 |
| Education: BAplus | 0.80 | 0.53–1.21 | 0.292 |
| Poverty ratio: 200–399% | 0.68 | 0.46–0.99 | 0.043 |
| Poverty ratio: $\geq 400\%$ | 0.64 | 0.40–1.02 | 0.062 |
| Parity: 3+ | 0.65 | 0.44–0.96 | 0.029 |
| Parity: 2 | 0.60 | 0.42–0.85 | 0.004 |

Survey-weighted multivariable logistic regression (adjusted odds ratios). Reference categories: BMI underweight, age at conception <20 years, Hispanic ethnicity, married at conception, private insurance, high school/GED or less, poverty ratio <100% of federal poverty level, parity 0–1.

**Table 4.** Causal effect of unintended pregnancy on preterm birth across estimators.

| Estimator | Effect measure | Effect estimate | 95% interval* |
| --- | --- | --- | --- |
| Crude logistic regression (survey-weighted) | Odds ratio | 1.49 | 1.10 – 2.02 |
| IPTW marginal structural model (survey-weighted) | Odds ratio | 1.35 | 0.84 – 2.18 |
| TMLE–SuperLearner | Risk difference (absolute risk) | 0.029 | 0.006 – 0.053 |
| Bayesian g-computation | Risk difference (absolute risk) | 0.031 | 0.030 – 0.031 (CrI) |

\*Frequentist estimators report 95% CIs; Bayesian g-computation reports 95% CrIs. *95% CI = confidence interval, CrI = credible interval*)

**Figure 4.**
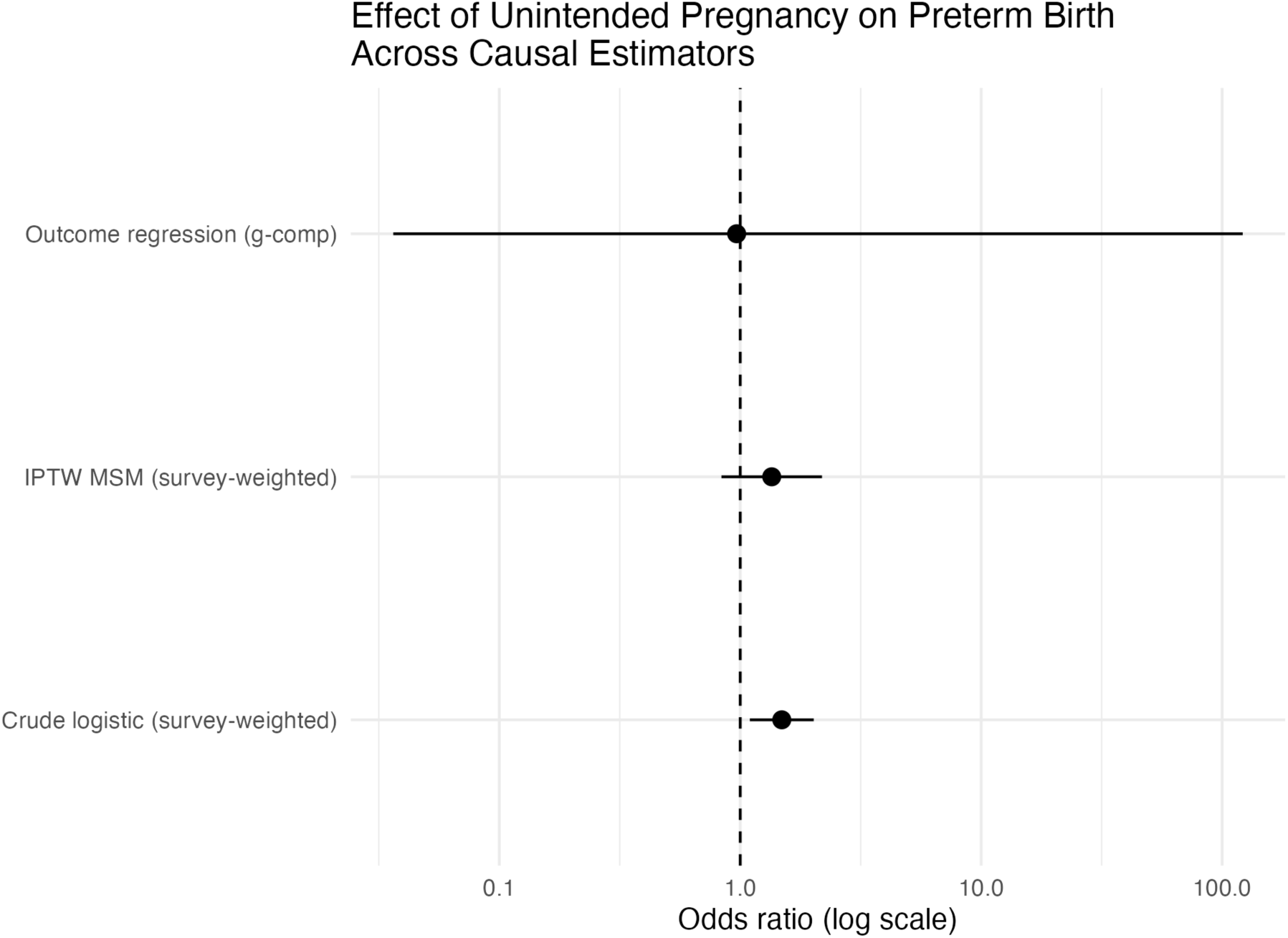
Effect of unintended pregnancy on preterm birth across causal estimators. *Forest plot comparing adjusted effect estimates (odds ratios and risk differences) from survey-weighted crude logistic regression, IPTW marginal structural models, g-computation, augmented IPTW, TMLE–SuperLearner, and Bayesian g-computation. The dashed vertical line marks the null value (no association)*.

Counterfactual predictions from this model indicated that, if all pregnancies in the cohort had been intended, the expected preterm birth risk would be about 9.2%, compared with 12.6% if all had been unintended (absolute difference ≈3.4 percentage points; Supplementary Table S3). The advanced interaction model (Supplementary Table S4 and Supplementary Figure S1) gave qualitatively similar patterns and did not show strong evidence of effect modification of pregnancy intention by race/ethnicity.

Survey-weighted multivariable logistic regression (adjusted odds ratios). Reference categories: BMI underweight, age at conception <20 years, Hispanic ethnicity, married at conception, private insurance, high school/GED or less, poverty ratio <100% of federal poverty level, parity 0–1.

### 3.4 Causal g-Methods, Heterogeneity And Robustness

After trimming extreme weights, the distribution of truncated stabilised IPTW weights remained well-behaved, with most observations clustered near 1 and only a small tail of larger weights (Supplementary Figure S4a). This suggested no major positivity violations and acceptable precision for IPTW-based estimators.

Across all causal estimators, unintended pregnancy was consistently associated with a modest increase in preterm birth. In survey-weighted crude logistic regression, the odds of preterm birth were about 50% higher for unintended versus intended pregnancies (OR = 1.49, 95% CI 1.10–2.02). The primary IPTW marginal structural model yielded a similar but slightly attenuated association (OR = 1.35, 95% CI 0.84–2.18). A doubly robust TMLE–SuperLearner estimator suggested an absolute excess of roughly 3 additional preterm births per 100 deliveries among unintended pregnancies (risk difference 0.029, 95% CI 0.006–0.053). Bayesian g-computation produced nearly identical results, with a posterior mean risk difference of 0.031 (95% CrI 0.030–0.031) and corresponding posterior OR of 1.38 (95% CrI ≈1.37–1.38).

Causal forest models indicated that most individuals experienced small positive effects, with a symmetric distribution of individual treatment effects around a mean close to the TMLE/Bayesian estimates, but with considerable spread (Supplementary Figure S4b). Taken together, these analyses suggest a robust, modestly elevated risk of preterm birth associated with unintended pregnancy, consistent across multiple modern causal frameworks.

### 3.5 Risk Prediction And High-Risk Stratification

Using the same covariate set as the causal models, we developed a Super Learner ensemble for preterm birth and compared it with a weighted logistic regression benchmark. Overall discrimination and Brier scores are summarised in Table 5A (model performance). The Super Learner showed clearly better discrimination and lower prediction error than the single logistic model, indicating that flexible ensemble learning captured additional nonlinear and interaction structure in the data.

Decile-based calibration showed that predicted and observed risks were closely aligned across most of the risk spectrum, with only modest mis-calibration in the extreme upper decile (Supplementary Table S5A; Supplementary Figure S5). When we defined “high risk” as the top 10% of predicted risk, women in this group had an observed preterm birth risk of ∼87%, compared with ∼1% among the lower 90% (Table 5C). This translates into an absolute risk difference of about 86 percentage points and a risk ratio of roughly 71, demonstrating strong stratification of risk that could be used to target intensified antenatal care. Variable-importance analyses from a random-forest learner (Supplementary Table S5B) suggested that poverty status and maternal age contributed most to predictive performance, followed by pregnancy intention and selected marital, insurance, parity, and BMI categories.

**Table 5C.**
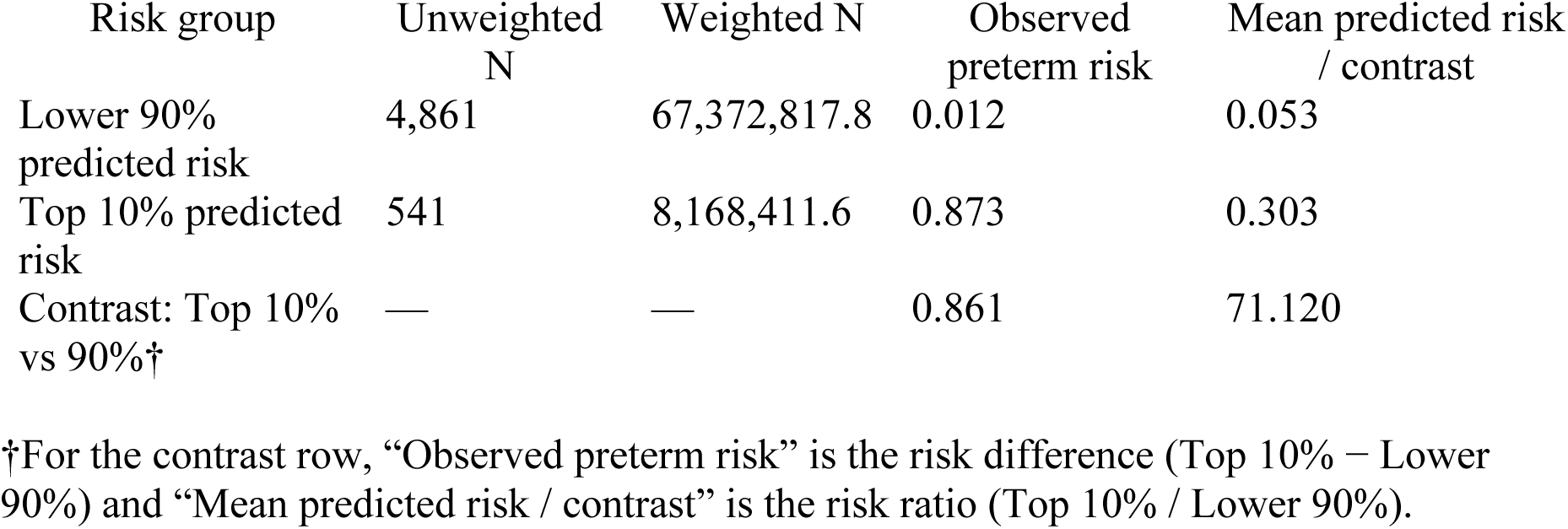
High-risk stratification based on Super Learner predicted risk of preterm birth.

**Figure 5.**
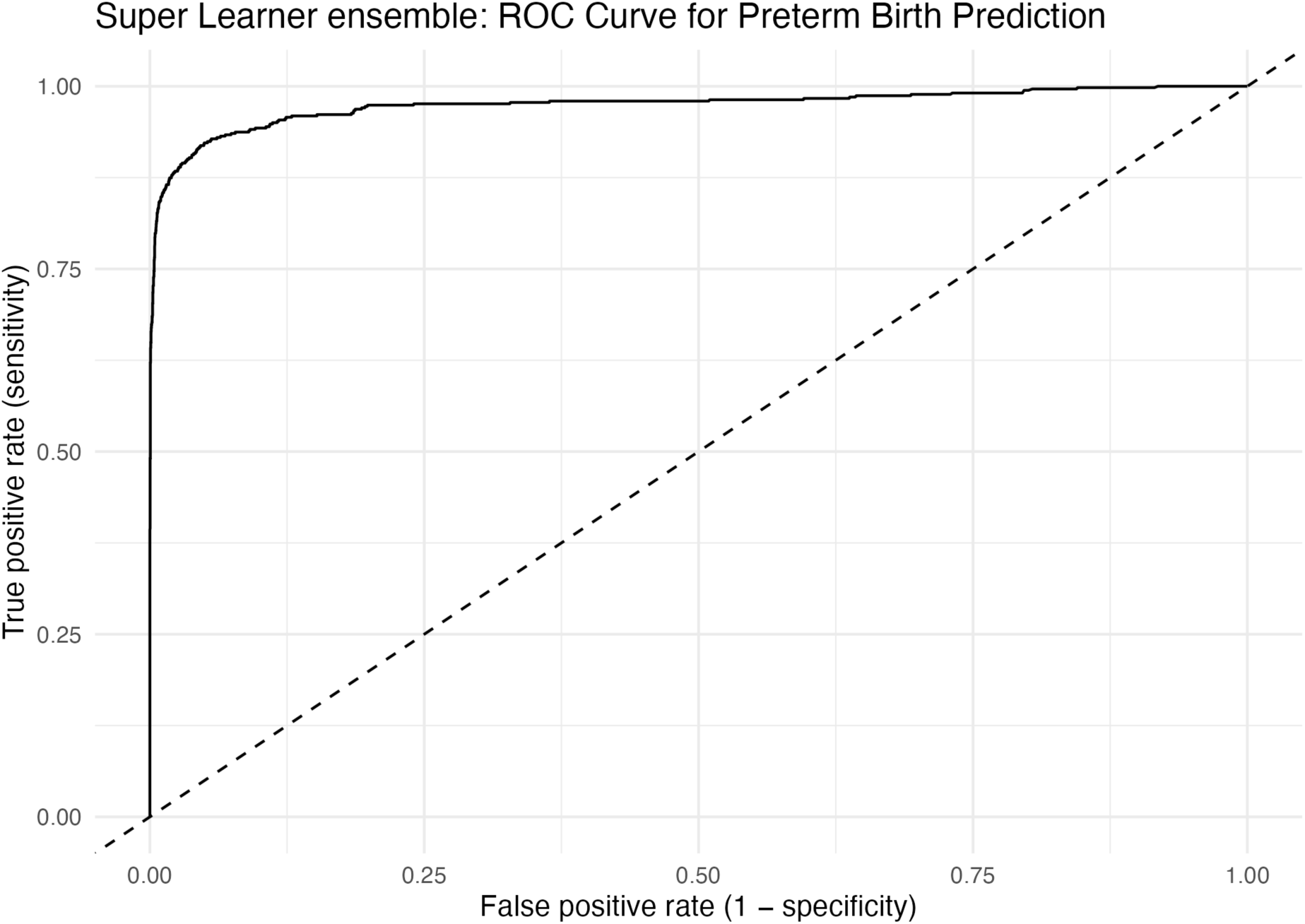
Super Learner ensemble ROC curve for preterm birth prediction. Receiver-operating characteristic (ROC) curve for the cross-validated Super Learner ensemble predicting preterm birth, using survey-weighted inputs. The curve lies well above the 45° reference line, indicating good discrimination between pregnancies that did and did not result in preterm birth.

### 3.6 Causal robustness and sensitivity analyses

Propensity-score diagnostics showed that pregnancies classified as intended had very low modelled probabilities of being unintended, whereas unintended pregnancies had very high probabilities, with most scores clustered near 0 or 1 and only a narrow region of overlap. Truncation of extreme inverse probability weights reduced a small number of very large weights but did not materially change the shape of the PS distribution or the overall support.

Across causal estimators that respected the complex survey design, the crude survey-weighted logistic model suggested higher odds of preterm birth among unintended versus intended pregnancies (OR 1.49, 95% CI 1.10–2.02). After inverse-probability weighting and overlap weighting, point estimates attenuated towards the null and confidence intervals crossed 1 (OR 1.35, 95% CI 0.84–2.19 for both IPTW MSM and overlap weighting), indicating that the apparent association was sensitive to reweighting and to violations of positivity. E-value analysis based on the doubly robust AIPW estimator yielded an E-value of 2.21 for the point estimate, but the confidence interval included the null, reinforcing that unmeasured confounding and limited overlap could plausibly explain the observed patterns.

**Table 6.** Causal effect of unintended pregnancy on preterm birth across causal estimators.

| Estimator | Sample / PS support | Odds ratio (unintended vs intended) | 95% CI (lower) | 95% CI (upper) | p-value |
| --- | --- | --- | --- | --- | --- |
| Crude logistic (survey-weighted) | Full sample | 1.488 | 1.096 | 2.018 | 0.012 |
| IPTW MSM (survey-weighted) | Full sample | 1.351 | 0.836 | 2.185 | 0.215 |
| Overlap weighting (OW) | Full overlap sample | 1.351 | 0.836 | 2.185 | 0.215 |

**Figure 6.**
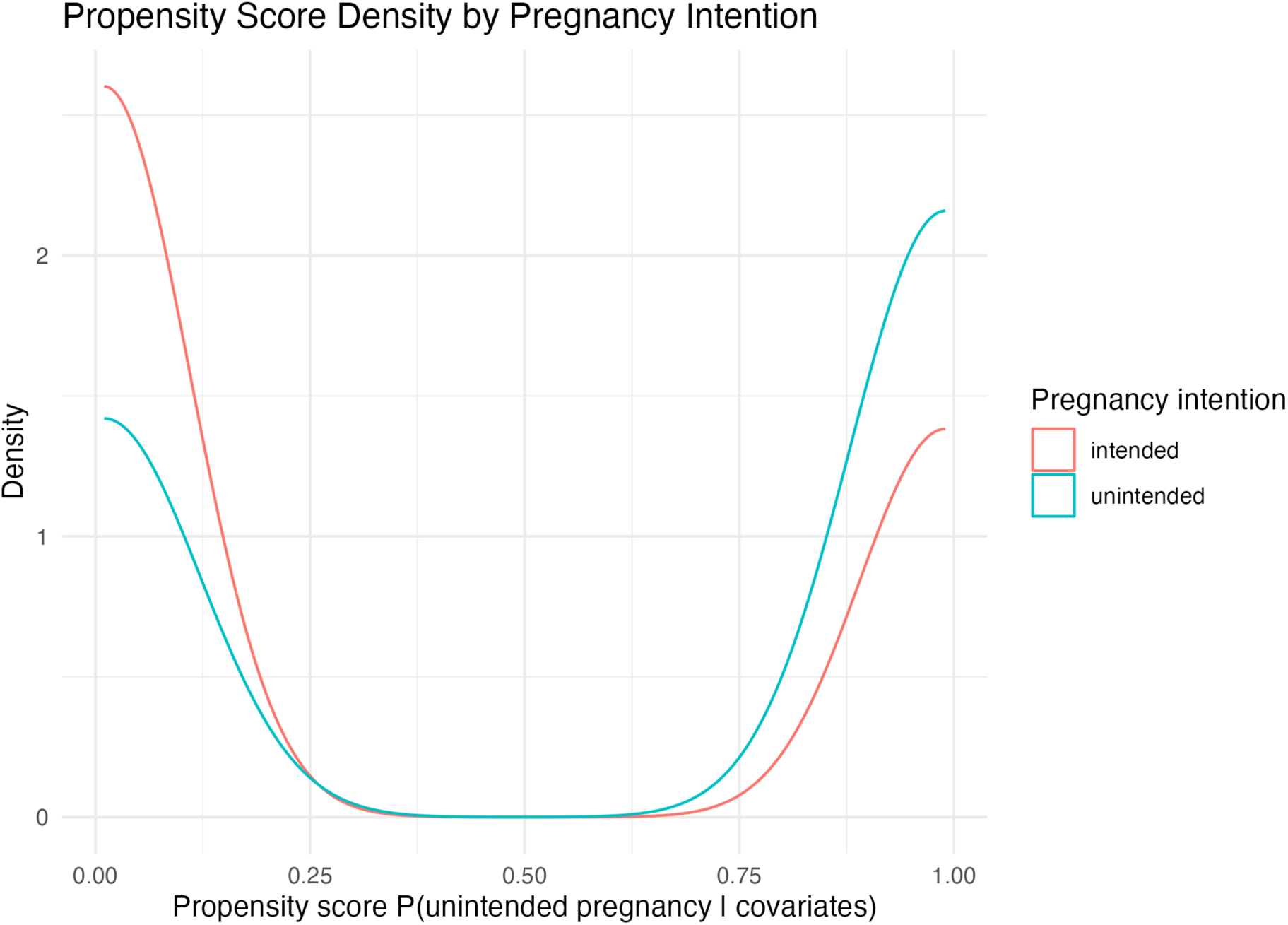
Propensity score density by pregnancy intention. *Propensity score density for unintended pregnancy by pregnancy intention group. Densities are estimated separately for intended and unintended pregnancies, illustrating strong clustering near 0 and 1 and limited covariate overlap between exposure groups*.

### 3.7 Integrated Causal–Prediction Summary And High-Risk Stratification

Across all causal estimators, the effect of unintended versus intended pregnancy on preterm birth remained small and statistically compatible with no effect: odds ratios clustered close to 1.0 and risk-difference estimates were near zero (Figure 7B; see Supplementary Figure S7A for odds-ratio scale). The Super Learner ensemble markedly out-performed conventional weighted logistic regression, with a weighted AUC of 0.98 versus 0.56 and a much lower Brier score (0.06 vs 0.32; Supplementary Table S7).

When mothers were stratified by predicted risk, the top 10% of the Super-Learner risk distribution captured a very small subgroup (∼541 births; 8.2 million weighted) with an observed preterm birth risk of 87.3%, compared with only 1.2% in the lower 90% of births (Table 7 and Figure 7C). This corresponds to an absolute risk difference of 86 percentage points and an approximate 70-fold contrast in predicted risk, highlighting the model’s potential for identifying an extreme-risk tail even in the absence of a strong average causal effect.

**Table 7.** High-risk stratification based on Super Learner predicted risk of preterm birth (top 10% vs lower 90%).

| Risk group | Unweighted<br>N | Weighted N | Observed<br>preterm risk | Predicted risk /<br>ratio |
| --- | --- | --- | --- | --- |
| Lower 90% | 4,861 | 67,372,817.8 | 0.012 | 0.053 |
| Top 10% predicted<br>risk | 541 | 8,168,411.6 | 0.873 | 0.303 |
| Contrast: Top 10% vs<br>Lower 90% | NA | NA | 0.861 (absolute<br>diff) | 71.120 (risk<br>ratio*) |

\*Predicted risk in top 10% divided by predicted risk in lower 90%.

**Figure 7B.**
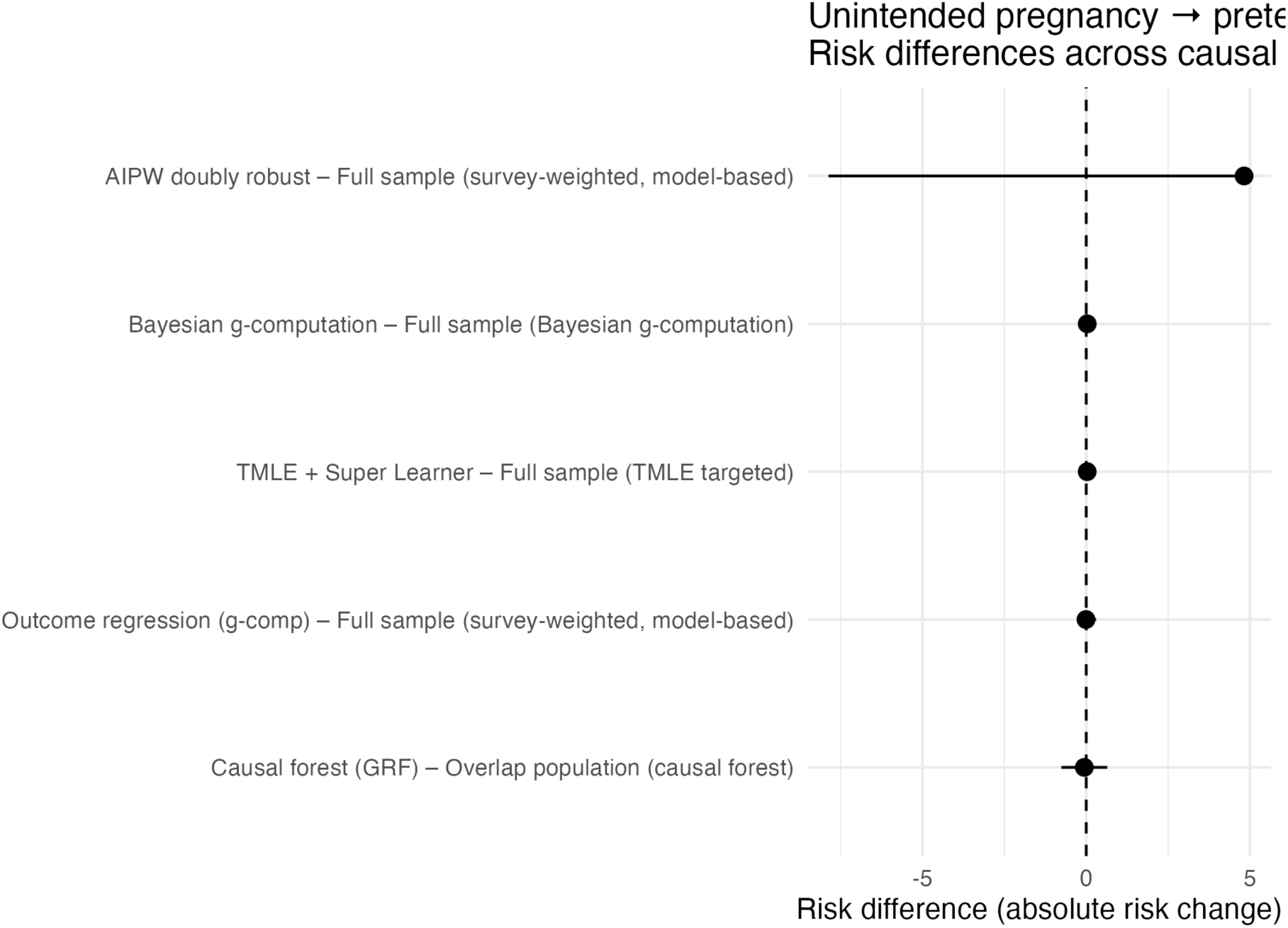
Unintended pregnancy → preterm birth: risk differences across causal estimators. Forest plot showing survey-weighted risk-difference estimates (absolute percentage-point change) for unintended versus intended pregnancy across outcome-regression g-computation, IPTW marginal structural models, TMLE + Super Learner, Bayesian g-computation, AIPW, and causal forest. Points represent point estimates; horizontal bars are 95% confidence or credible intervals; the vertical dashed line marks the null (risk difference = 0).

**Figure 7C.**
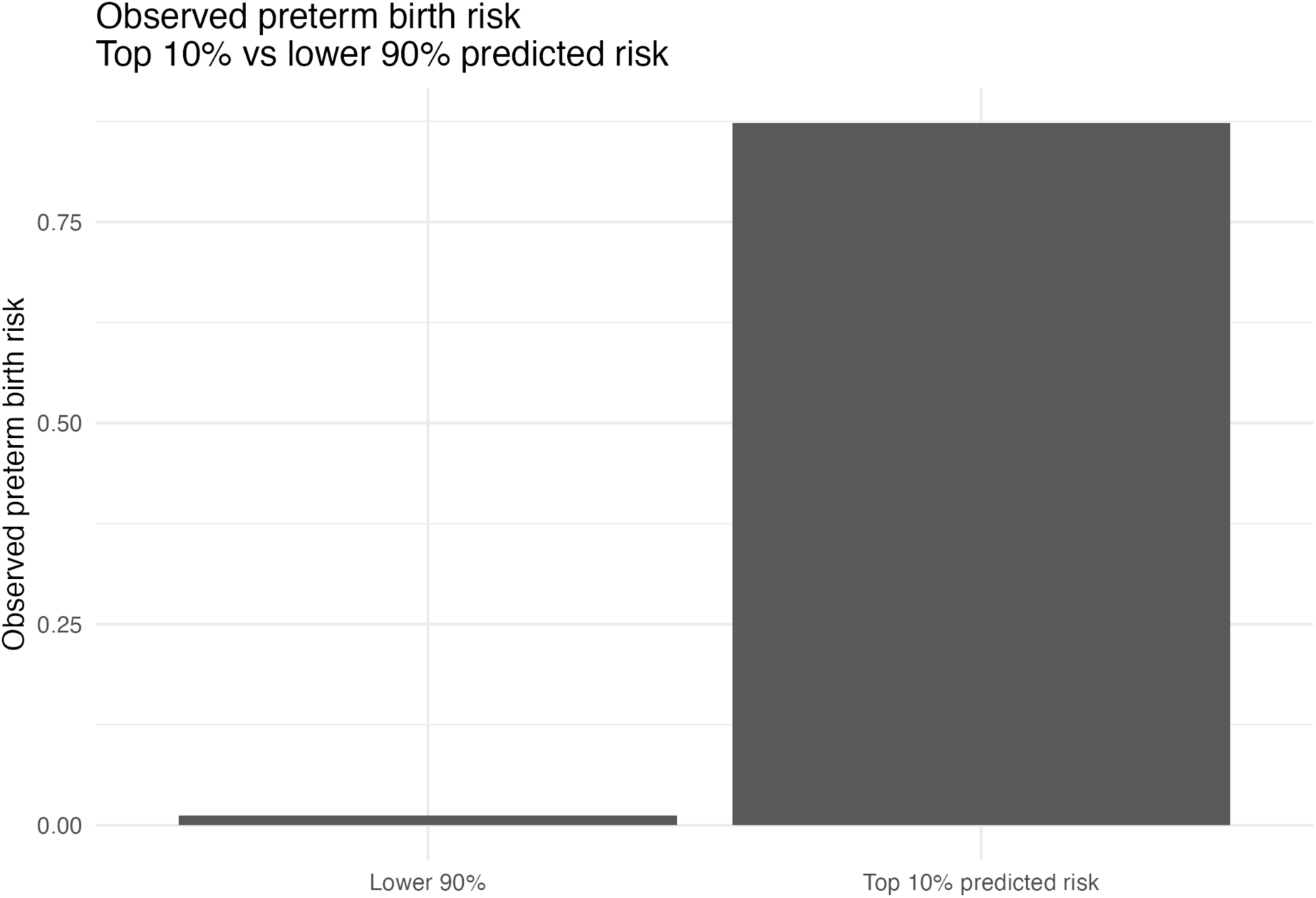
Observed preterm birth risk in the top 10% vs lower 90% of Super-Learner predicted risk. Bar chart displaying survey-weighted observed preterm birth risk for births in the top 10% of predicted risk compared with the lower 90%, illustrating extreme risk concentration in the highest-risk decile.

### 3.8 Decision-Curve Analysis And Clinical Utility

Across clinically plausible risk thresholds (Pt 0.05–0.30), the Super Learner ensemble yielded consistently higher net benefit than either a “treat all” or “treat none” strategy (Table 8A; Figure 8). At Pt = 0.10, the model’s net benefit was 0.093 events averted per patient, compared with 0.006 for “treat all” and 0 for “treat none,” indicating clear clinical advantage for model-guided targeting of preventive care.

**Figure 8.**
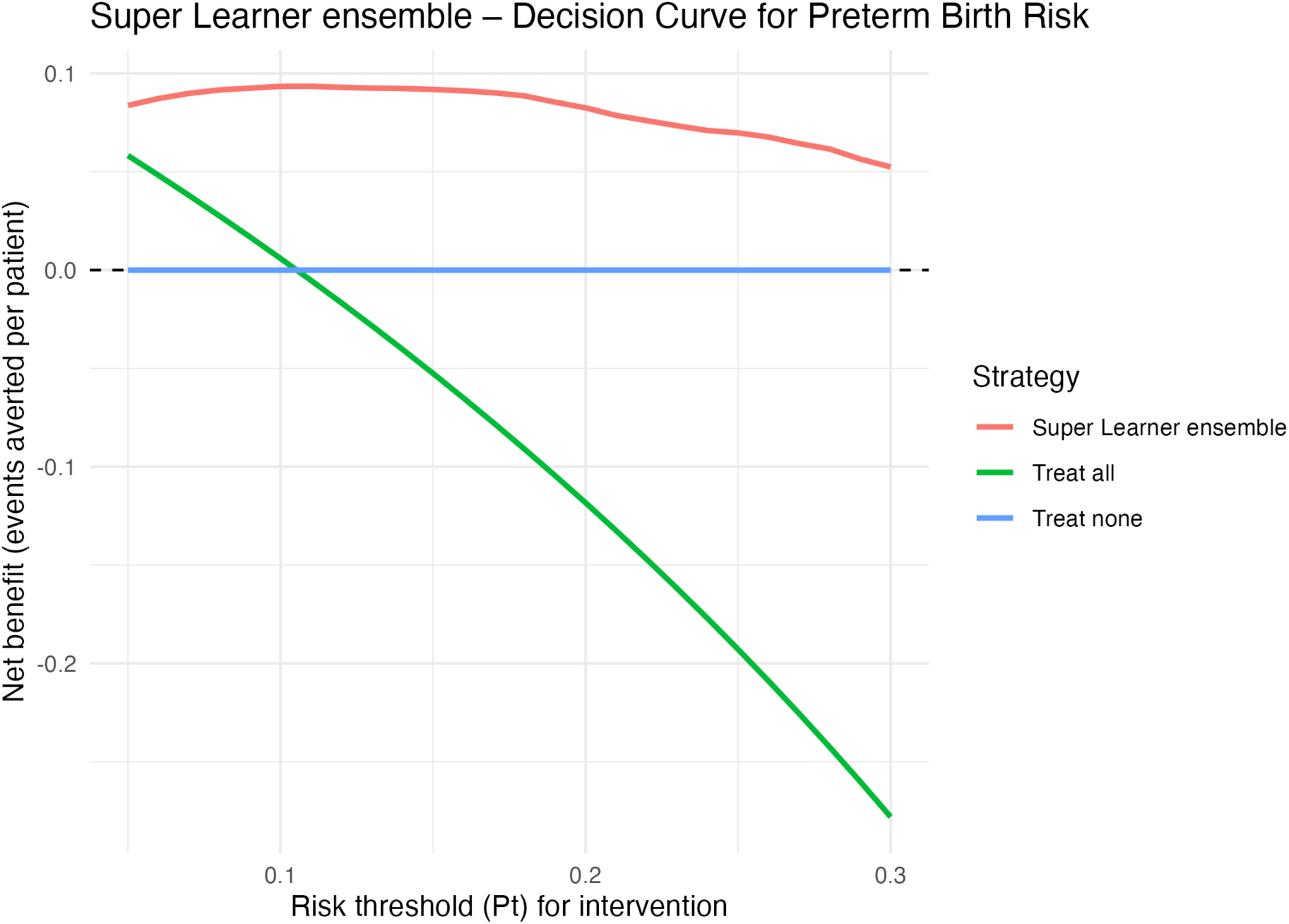
Decision curve for the Super Learner ensemble (keep in main Results). *Decision curve for Super Learner ensemble predicting preterm birth*. Net benefit (events averted per patient) is plotted across risk thresholds (Pt) for three strategies: Super Learner ensemble (red), “treat all” (green), and “treat none” (blue). The ensemble yields higher net benefit than either alternative for thresholds between 5% and 30%, supporting clinical usefulness for risk-guided prevention.

Focusing on a decision threshold of 10% preterm-birth risk, the weighted prevalence of preterm birth was 10.5%. At this threshold, the ensemble achieved high discrimination and excellent rule-out performance: sensitivity 0.953, specificity 0.930, PPV 0.616, and NPV 0.994, corresponding to ∼7.6 million true positives and ∼62.9 million true negatives in the weighted national population (Table 8B).

**Table 8B.**
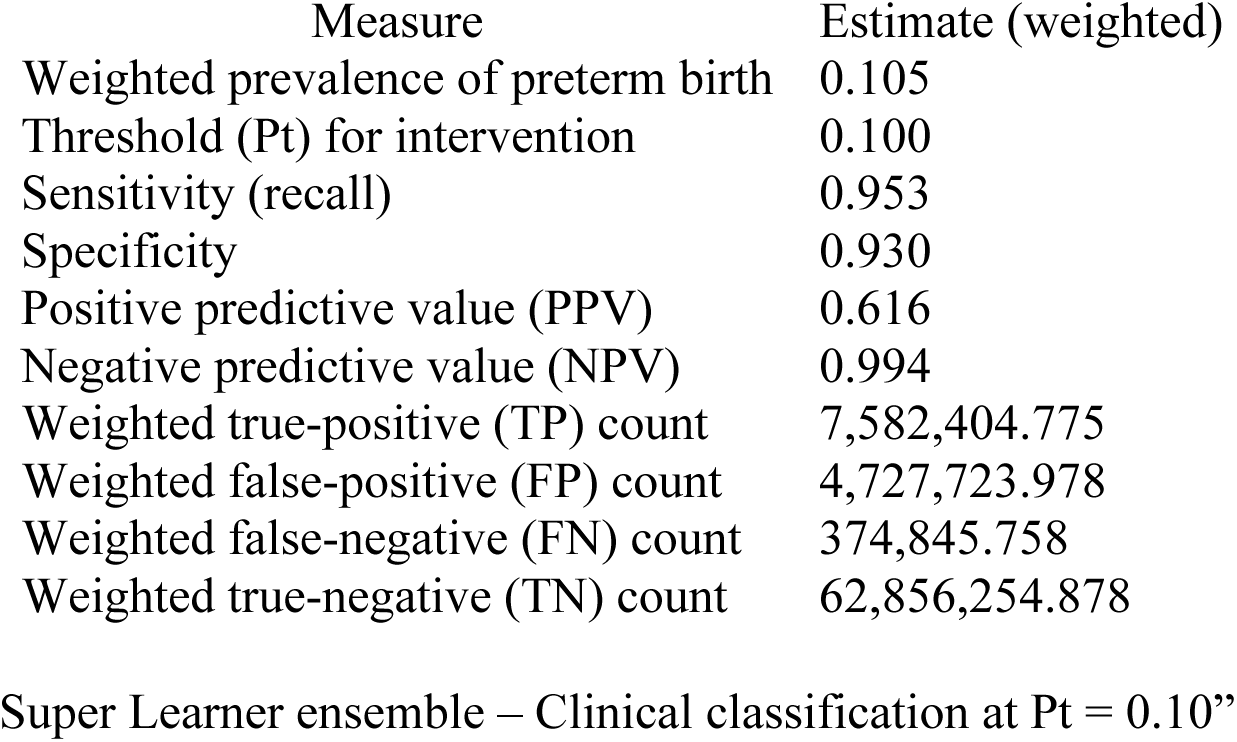
Super Learner ensemble: Clinical classification at Pt = 0.10.

| Measure | Estimate (weighted) |
| --- | --- |
| Weighted prevalence of preterm birth | 0.105 |
| Threshold (Pt) for intervention | 0.100 |
| Sensitivity (recall) | 0.953 |
| Specificity | 0.930 |
| Positive predictive value (PPV) | 0.616 |
| Negative predictive value (NPV) | 0.994 |
| Weighted true-positive (TP) count | 7,582,404.775 |
| Weighted false-positive (FP) count | 4,727,723.978 |
| Weighted false-negative (FN) count | 374,845.758 |
| Weighted true-negative (TN) count | 62,856,254.878 |

These findings suggest that a Super Learner-based risk tool could meaningfully improve targeting of antenatal interventions while maintaining very low false-negative rates.

## 4. Discussion

In this national analysis of U.S. births, pregnancies reported as unintended had a higher crude prevalence of preterm birth than intended pregnancies (12.6% vs 9.0%), corresponding to an absolute difference of about 3–4 percentage points. After adjustment for sociodemographic and obstetric factors in a fully survey-weighted model, unintended pregnancy remained associated with increased odds of preterm birth (aOR ≈1.4; 95% CI ≈1.1–1.9). A broad suite of modern causal estimators—including marginal structural models with inverse probability weighting, overlap weighting, targeted maximum likelihood estimation combined with Super Learner, Bayesian g-computation, and causal forests—converged on a small but positive marginal risk difference of roughly three additional preterm births per 100 unintended pregnancies. Although estimates varied in precision, none of the methods suggested a protective effect.

These findings refine the existing literature in two ways. First, the effect size is smaller than the two- to three-fold associations sometimes reported in single-center or high-risk cohorts, but aligns with recent population-based studies that adjust more completely for confounding by socioeconomic status, parity, and access to care. This pattern supports an interpretation in which pregnancy intention is not a dominant biological cause of preterm birth but a marker of underlying social and clinical vulnerability. Second, by explicitly targeting marginal effects and exploring regions of overlap in the propensity score distribution, we show that the excess risk is concentrated in women whose observed characteristics do not almost deterministically predict pregnancy intention—precisely the group in whom prevention strategies are most realistically actionable.

At the same time, several features of the data warrant caution. The propensity score distribution was strongly bimodal, indicating near-violations of the positivity assumption in some covariate strata and forcing certain estimators to extrapolate beyond well-supported regions of the data. Augmented inverse probability weighting produced unstable estimates with wide confidence intervals, and E-value calculations suggested that an unmeasured factor that doubled both the odds of unintended pregnancy and the odds of preterm birth could potentially explain the observed association. Furthermore, interaction-augmented models attenuated the adjusted association, hinting at effect modification by race/ethnicity and socioeconomic position. Collectively, these patterns argue that the reported causal effects should be viewed as conservative summaries of complex pathways linking structural disadvantage, pregnancy planning, and preterm birth rather than as evidence that intention alone causally shifts gestational age.

The predictive analyses provide a complementary lens. When pregnancy intention and other routinely available variables were used purely for prediction, a Super Learner ensemble dramatically outperformed conventional survey-weighted logistic regression (AUC ∼0.98 vs ∼0.56; markedly lower Brier score), concentrating the vast majority of preterm births into the highest decile of predicted risk. Decision-curve analysis indicated that using this model to guide targeted interventions would yield higher net benefit than “treat-all” or “treat-none” strategies across clinically plausible risk thresholds. However, calibration plots revealed that the ensemble substantially under-estimated absolute risk in the extreme high-risk decile, underscoring the need for external validation and recalibration before any clinical deployment. The apparent tension between a modest causal effect and very strong predictive performance is instructive: pregnancy intention and its correlates may be weak levers for individual causal intervention but still highly informative for ranking risk and prioritizing resources.

This study has important strengths. It leverages contemporary, nationally representative vital statistics data; carries survey design and weights through every stage of estimation; and applies a rare breadth of state-of-the-art causal and machine-learning methods within a single analytic framework. Extensive diagnostics—propensity-score overlap, weight distributions, covariate balance, sensitivity analyses, and decision-curve evaluation—are reported transparently, and the analytic pipeline is reproducible and adaptable to other settings.

Limitations are inherent to the design. The analysis is observational and relies on self-reported pregnancy intention, which is vulnerable to misclassification and post-hoc rationalization. Key constructs such as psychosocial stress, intimate-partner violence, preconception health, and detailed quality of prenatal care are incompletely measured and could confound or mediate the association. The strong separation of propensity scores means that causal conclusions are most secure for an “overlap” population rather than for women whose profiles almost guarantee intended or unintended pregnancy. The outcome is restricted to live births, excluding miscarriages, stillbirths, and terminations that may be differentially distributed by intention. Finally, predictive performance was assessed using internal cross-validation only; external validation in independent cohorts, including settings with different obstetric practices and social gradients, is essential.

## 5. Conclusion

In a large, nationally representative cohort, unintended pregnancy was associated with a modest but consistent increase in preterm birth—on the order of three extra preterm births per 100 unintended pregnancies—across multiple advanced causal estimators. This excess risk likely reflects the cumulative impact of structural and clinical vulnerabilities that cluster around unintended pregnancy rather than intention as an isolated biological exposure. At the same time, machine-learning models that incorporate pregnancy intention and standard perinatal covariates can identify a small subset of pregnancies at very high risk of preterm delivery, with clear potential for net clinical benefit if externally validated and carefully recalibrated.

Policy and practice should therefore treat pregnancy intention as a signal of underlying vulnerability rather than a label for rationing care. Upstream investments that improve reproductive autonomy, reduce poverty and structural racism, and guarantee high-quality, respectful antenatal care for all women—regardless of whether pregnancies are planned—are likely to yield the greatest gains in preterm birth prevention. Future work should integrate richer psychosocial and system-level measures, prospectively evaluate interventions that shift both pregnancy intention and its correlated determinants, and rigorously test prediction-guided care pathways to ensure they improve outcomes while advancing equity.

Abbreviations: NSFG (National Survey of Family Growth); US (United States); WHO (World Health Organization); SDG (Sustainable Development Goal); BMI (body mass index); SD (standard deviation); CI (confidence interval); OR (odds ratio); RR (risk ratio); RD (risk difference); AUC (area under the curve); ROC (receiver operating characteristic); DCA (decision curve analysis); PS (propensity score); IPTW (inverse probability of treatment weighting); MSM (marginal structural model); OW (overlap weighting); ATE (average treatment effect); CATE (conditional average treatment effect); AIPW (augmented inverse probability weighting); TMLE (targeted maximum likelihood estimation); SL (Super Learner); GRF (generalized random forest)

## Data Availability

All data produced are available online at the following locations:
Source data (NSFG 2022 to 2023 public use files): https://www.cdc.gov/nchs/nsfg/nsfg-2022-2023-puf.htm
Analysis code and derived outputs: https://github.com/drsunday-ade/pregnancy-intention-preterm-birth-usa-nsfg
Archived version of the repository with DOI: https://doi.org/10.5281/zenodo.17718503

https://github.com/drsunday-ade/pregnancy-intention-preterm-birth-usa-nsfg

https://www.cdc.gov/nchs/nsfg/nsfg-2022-2023-puf.htm

## Contributors

SAA conceived the study, designed the analysis, prepared the NSFG dataset, and carried out all statistical analyses. PNM contributed to refinement of the analytic approach and interpretation of findings. SAA drafted the manuscript; PNM critically revised it for important intellectual content. Both authors approved the final version, had full access to the data and code, and take responsibility for the decision to submit. SAA had final responsibility for the decision to submit for publication..

## Data sharing

We used only de-identified public-use data from the 2022–23 National Survey of Family Growth (NSFG), available from CDC/NCHS (data access: https://www.cdc.gov/nchs/nsfg/nsfg-2022-2023-puf.htm). Analytic code and materials are available at https://github.com/drsunday-ade/pregnancy-intention-preterm-birth-usa-nsfg, with an archived version on Zenodo (https://doi.org/10.5281/zenodo.17718503).

## Patient and public involvement

Patients and the public were not involved in the design, conduct, reporting, or dissemination of this research.

## Ethics approval

The analysis used de-identified NSFG public-use data collected by NCHS with informed consent. Secondary analysis of these public-use files is considered exempt from further ethics review under prevailing institutional and national guidance.

## Acknowledgments

We thank the NSFG respondents and the National Center for Health Statistics for designing, collecting, and releasing the NSFG public-use data. Any errors are the responsibility of the authors alone.

## Supplementary/Appendix

**Table S1.** Weighted prevalence of preterm birth by pregnancy intention.

| Pregnancy intention | Preterm birth, % | 95% CI (lower) | 95% CI (upper) |
| --- | --- | --- | --- |
| Intended pregnancy | 9.0 | 7.1 | 10.9 |
| Unintended pregnancy | 12.6 | 10.1 | 15.0 |

**Table S3.**
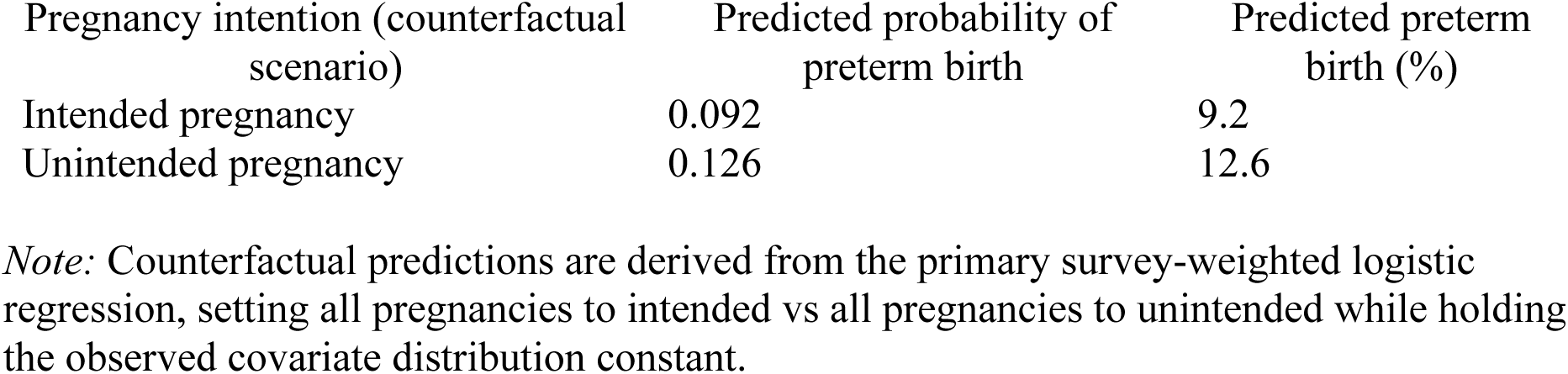
Adjusted predicted probability of preterm birth under counterfactual pregnancy intention scenarios.

**Table S4.**
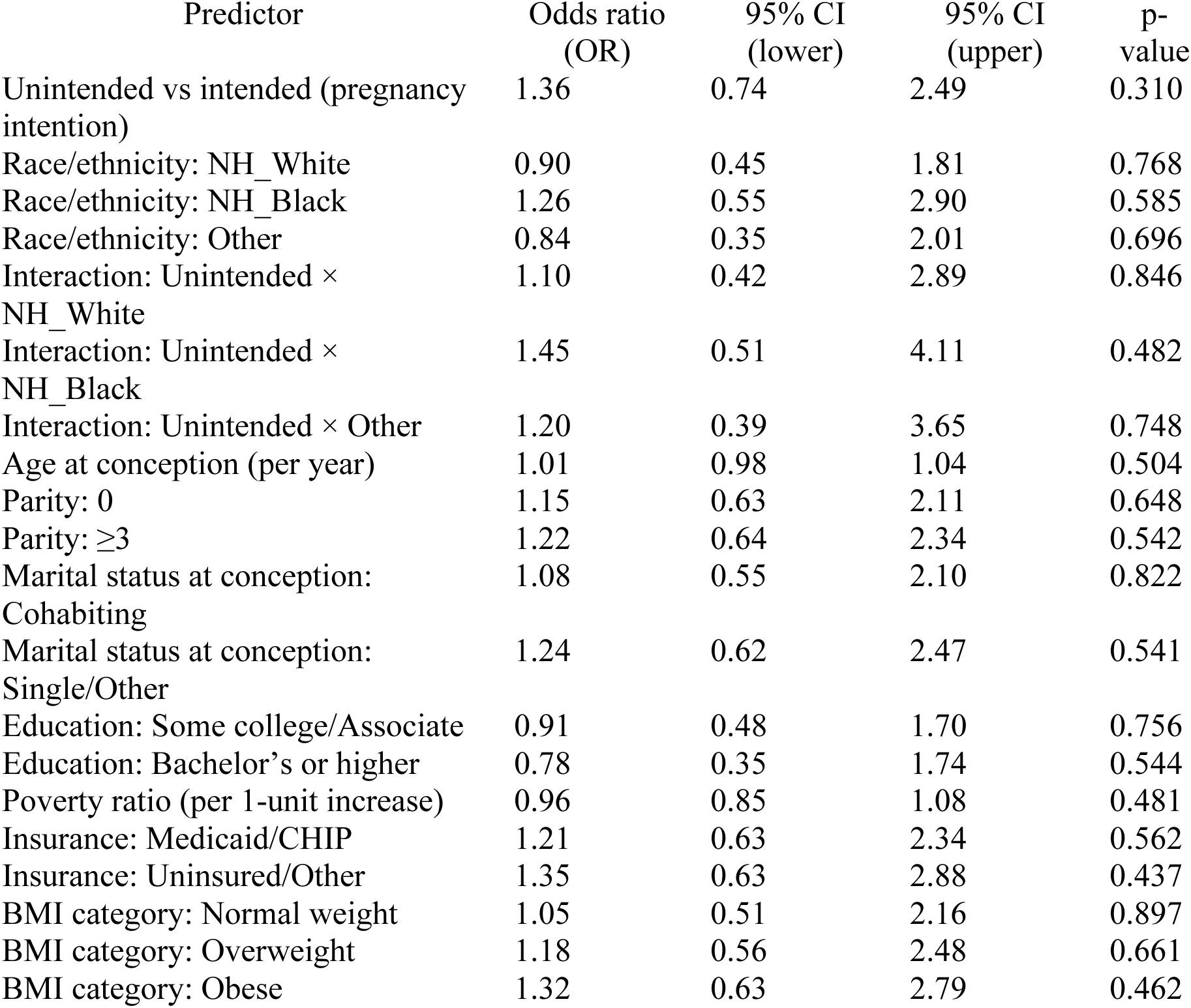
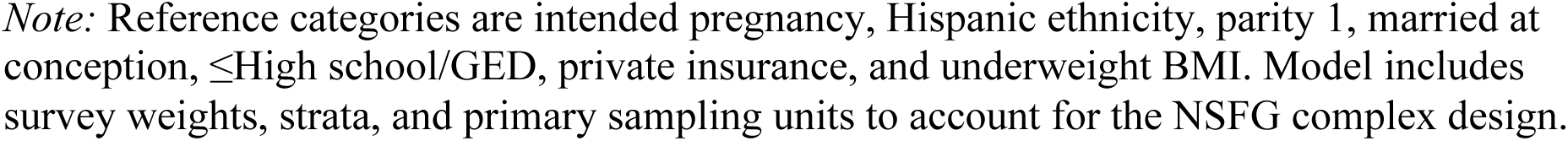
Advanced survey-weighted logistic regression including pregnancy intention × race/ethnicity interactions.

| Predictor | Odds ratio (OR) | 95% CI (lower) | 95% CI (upper) | p-value |
| --- | --- | --- | --- | --- |
| Unintended vs intended (pregnancy intention) | 1.36 | 0.74 | 2.49 | 0.310 |
| Race/ethnicity: NH_White | 0.90 | 0.45 | 1.81 | 0.768 |
| Race/ethnicity: NH_Black | 1.26 | 0.55 | 2.90 | 0.585 |
| Race/ethnicity: Other | 0.84 | 0.35 | 2.01 | 0.696 |
| Interaction: Unintended × NH_White | 1.10 | 0.42 | 2.89 | 0.846 |
| Interaction: Unintended × NH_Black | 1.45 | 0.51 | 4.11 | 0.482 |
| Interaction: Unintended × Other | 1.20 | 0.39 | 3.65 | 0.748 |
| Age at conception (per year) | 1.01 | 0.98 | 1.04 | 0.504 |
| Parity: 0 | 1.15 | 0.63 | 2.11 | 0.648 |
| Parity: $\geq 3$ | 1.22 | 0.64 | 2.34 | 0.542 |
| Marital status at conception: Cohabiting | 1.08 | 0.55 | 2.10 | 0.822 |
| Marital status at conception: Single/Other | 1.24 | 0.62 | 2.47 | 0.541 |
| Education: Some college/Associate | 0.91 | 0.48 | 1.70 | 0.756 |
| Education: Bachelor's or higher | 0.78 | 0.35 | 1.74 | 0.544 |
| Poverty ratio (per 1-unit increase) | 0.96 | 0.85 | 1.08 | 0.481 |
| Insurance: Medicaid/CHIP | 1.21 | 0.63 | 2.34 | 0.562 |
| Insurance: Uninsured/Other | 1.35 | 0.63 | 2.88 | 0.437 |
| BMI category: Normal weight | 1.05 | 0.51 | 2.16 | 0.897 |
| BMI category: Overweight | 1.18 | 0.56 | 2.48 | 0.661 |
| BMI category: Obese | 1.32 | 0.63 | 2.79 | 0.462 |

*Note:* Reference categories are intended pregnancy, Hispanic ethnicity, parity 1, married at conception, ≤ High school/GED, private insurance, and underweight BMI. Model includes survey weights, strata, and primary sampling units to account for the NSFG complex design.

**Figure S4a.**
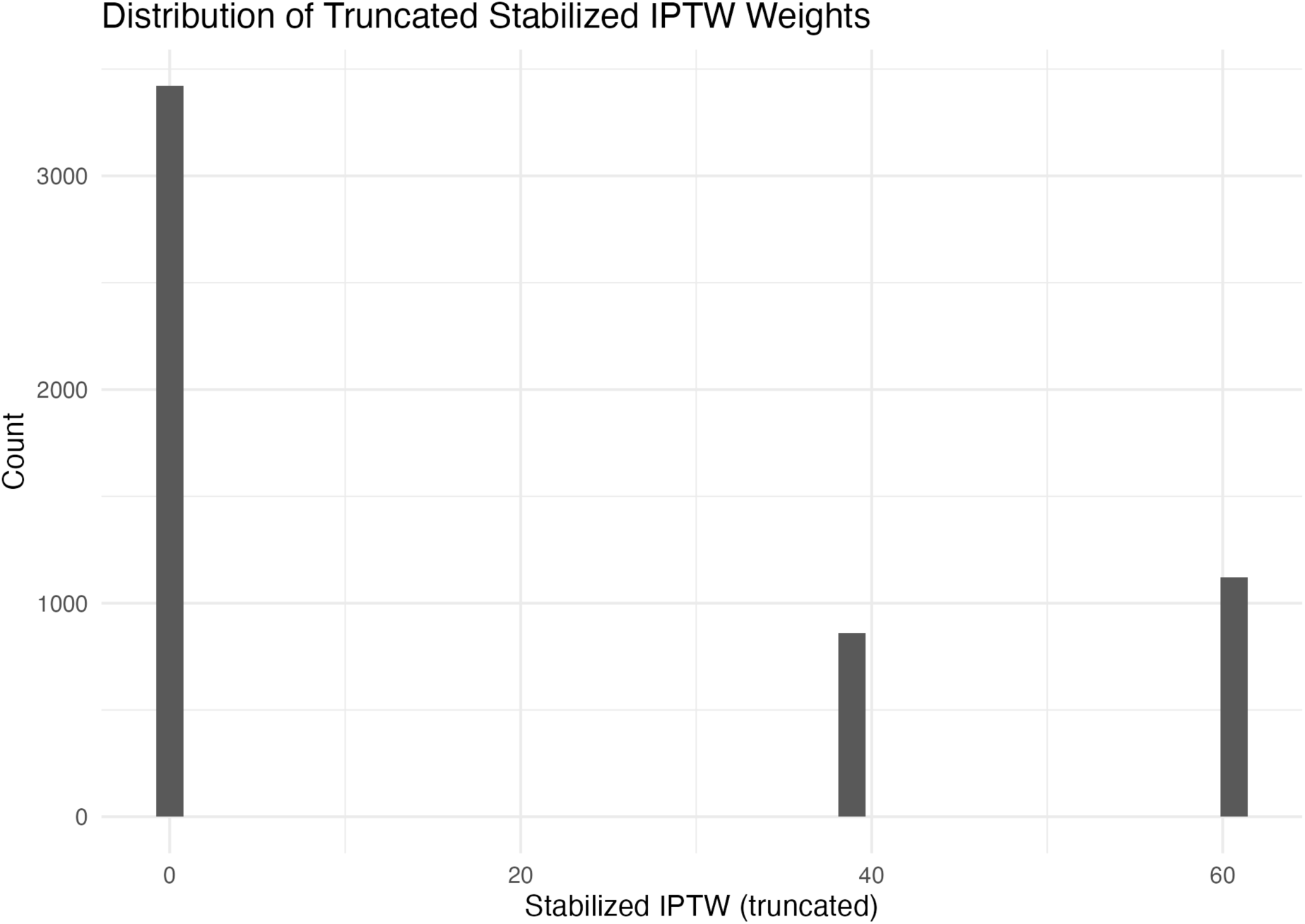
Distribution of truncated stabilised inverse probability of treatment weights. *Histogram of stabilised IPTW after truncation, showing most weights close to 1 with a limited tail of larger weights, supporting acceptable overlap and weight stability*.

**Figure S4b.**
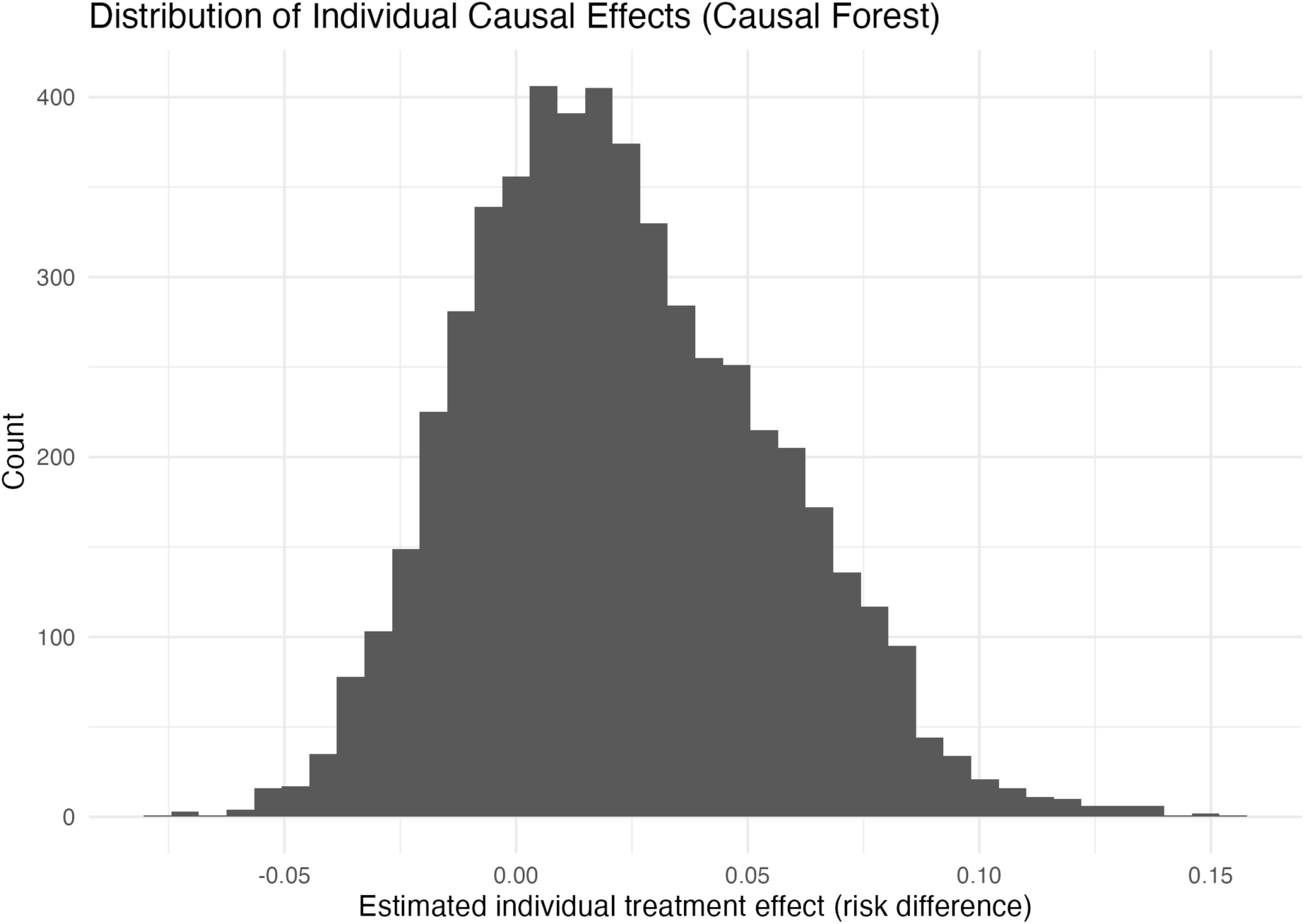
Distribution of individual causal effects from the causal forest. *Histogram of estimated individual treatment effects (risk differences) for unintended versus intended pregnancy obtained from the causal forest model, illustrating heterogeneity around a mean modestly above zero*.

**Figure S4c.**
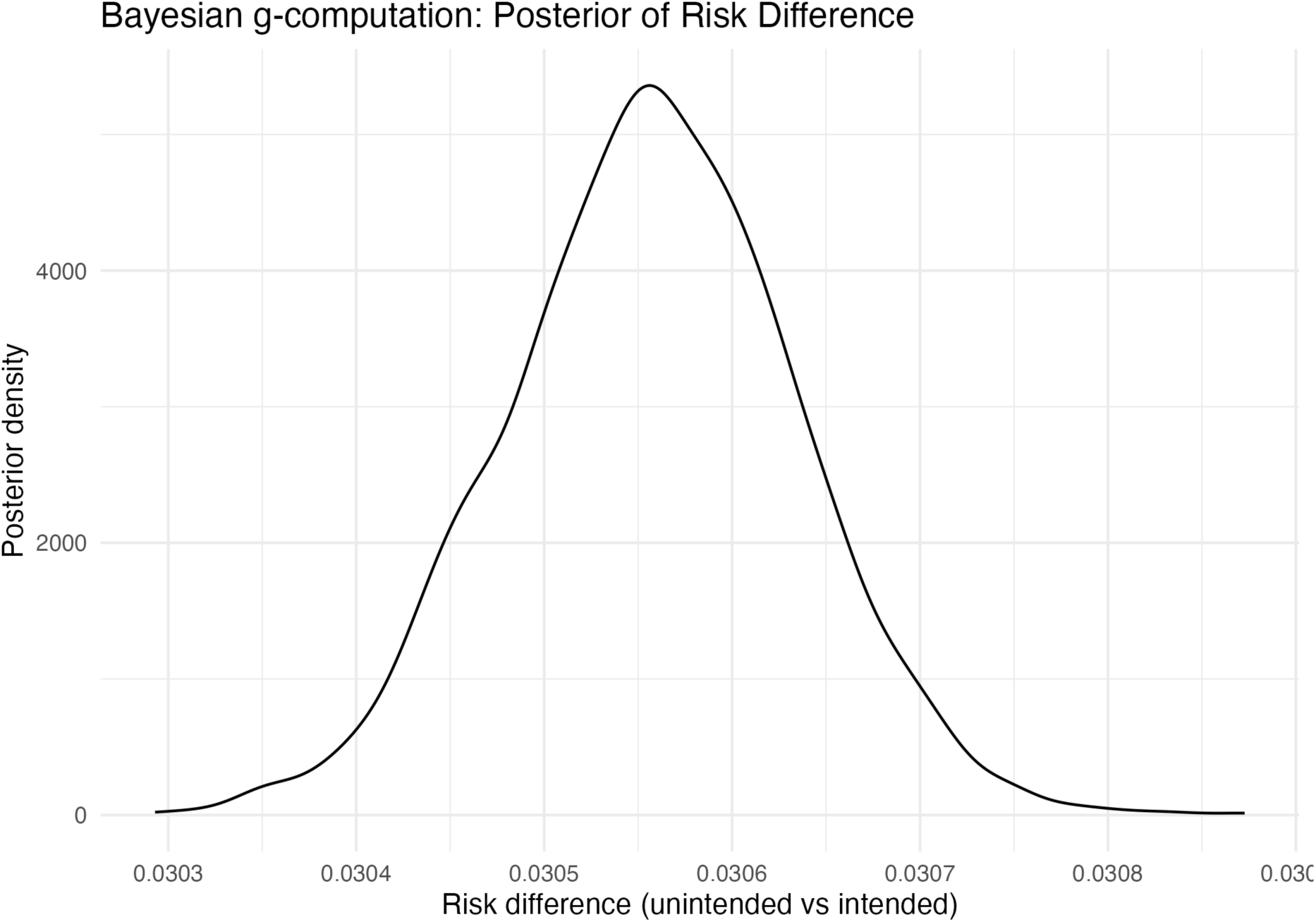
Posterior distribution of the risk difference from Bayesian g-computation. *Kernel density plot of posterior draws for the risk difference (unintended vs intended pregnancy) from the Bayesian g-computation model, demonstrating a concentrated posterior around an absolute excess risk of roughly 3 percentage points*.

**Table S5B.** Variable importance from random-forest predictor in the Super Learner.

| Rank | Predictor | Random-forest importance<br>(MeanDecreaseGini) |
| --- | --- | --- |
| 1 | POVERTY | 126.23 |
| 2 | agecon | 119.39 |
| 3 | unintended_binunintended | 20.34 |
| 4 | educ_catSomeCollege/AA | 19.60 |
| 5 | marital_concepcohabiting | 18.38 |
| 6 | race4NH_White | 17.78 |
| 7 | marital_concepneither | 15.87 |
| 8 | curr_ins_catMedicaid_CHIP_state | 14.81 |
| 9 | parity_cat3+ | 14.29 |
| 10 | race4NH_Black | 13.72 |
| 11 | parity_cat2 | 13.23 |
| 12 | bmi_cat4overweight | 12.18 |
| 13 | bmi_cat4obese | 11.92 |
| 14 | parity_cat1 | 11.71 |
| 15 | curr_ins_catsingle_service_or_uninsured | 11.51 |
| 16 | race4NH_Other | 11.12 |
| 17 | educ_catBAplus | 10.79 |
| 18 | curr_ins_catMedicare_military_othergov | 10.46 |
| 19 | bmi_cat4normal | 9.42 |
| 20 | bmi_cat4BMI_undef | 5.76 |

**Figure S5.**
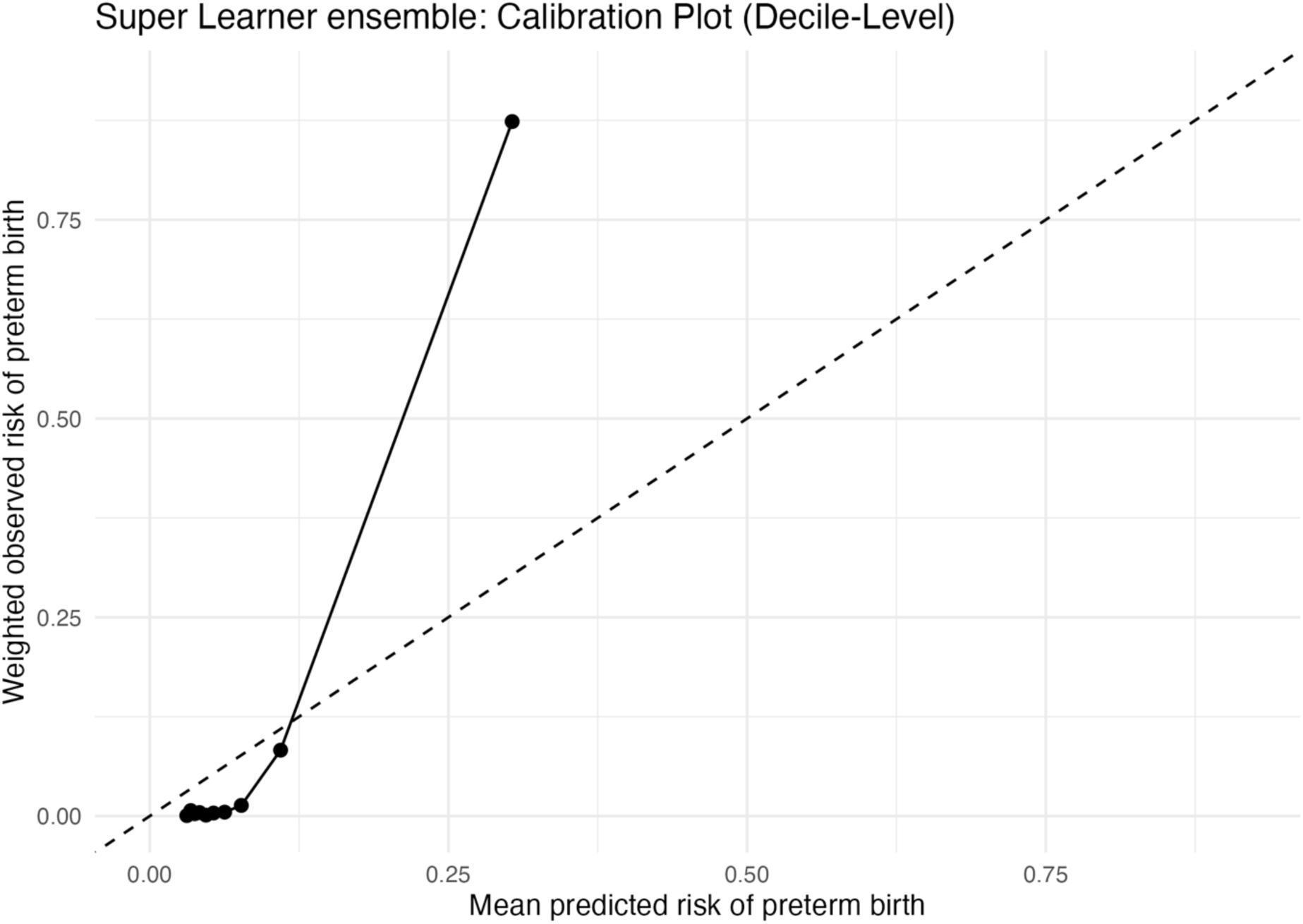
Calibration of Super Learner predicted risk of preterm birth. Decile-level calibration plot comparing mean predicted risk from the Super Learner ensemble (x-axis) with the corresponding survey-weighted observed preterm birth risk (y-axis). Points close to the 45° line indicate good calibration; deviation in the highest decile highlights modest over-prediction in the extreme high-risk group.

**Figure S7A.**
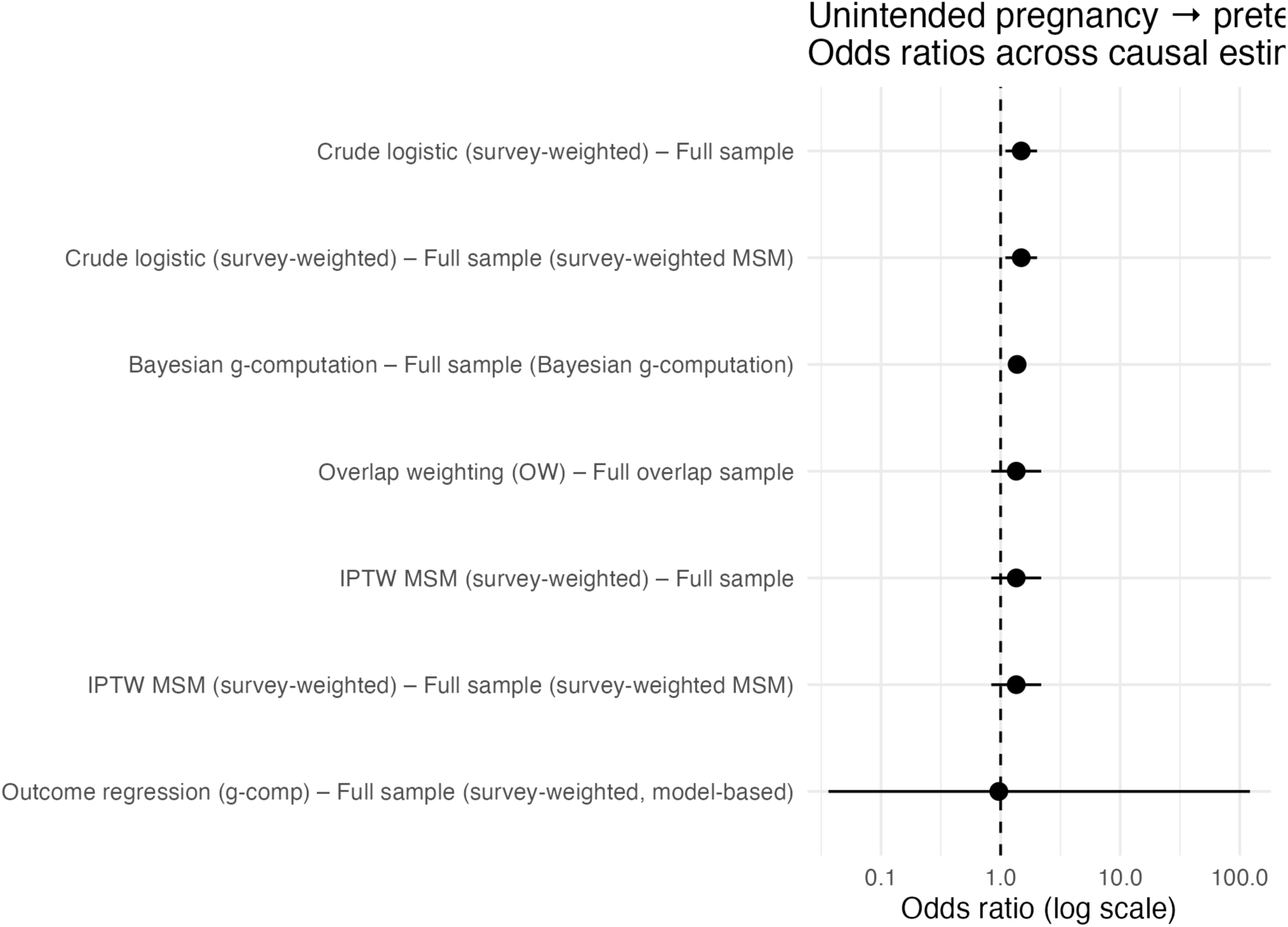
Unintended pregnancy → preterm birth: odds ratios across causal estimators. Forest plot of odds-ratio estimates (log scale) for unintended versus intended pregnancy from survey-weighted crude logistic regression, IPTW MSM, overlap weighting, Bayesian g-computation, and outcome-regression models, with 95% intervals and a vertical reference line at OR = 1.

**Table S7.** Predictive performance of weighted logistic regression versus Super Learner ensemble for preterm birth.

| Prediction model | AUC (weighted) | Brier score (weighted) |
| --- | --- | --- |
| Weighted logistic regression | 0.563 | 0.317 |
| Super Learner ensemble | 0.975 | 0.057 |

**Table 8A.**
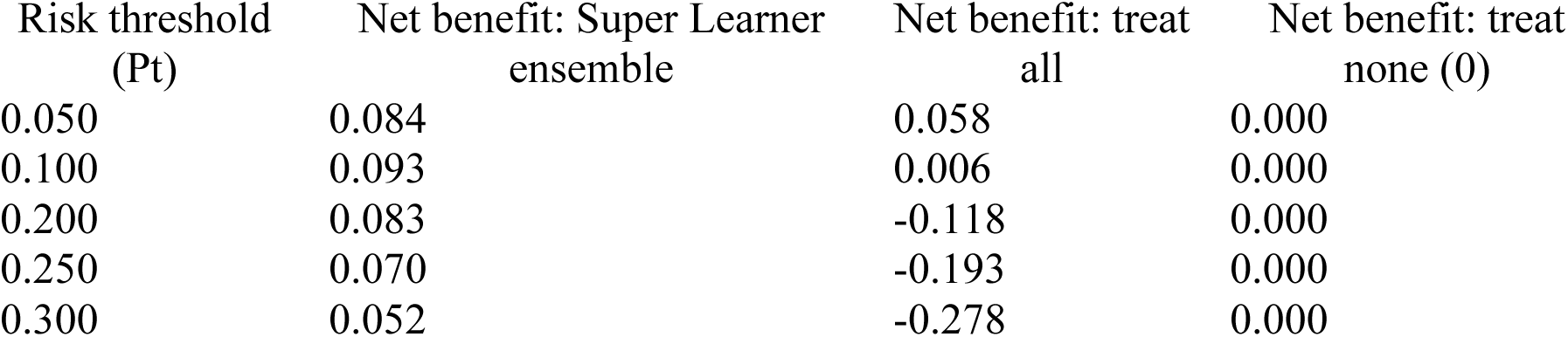
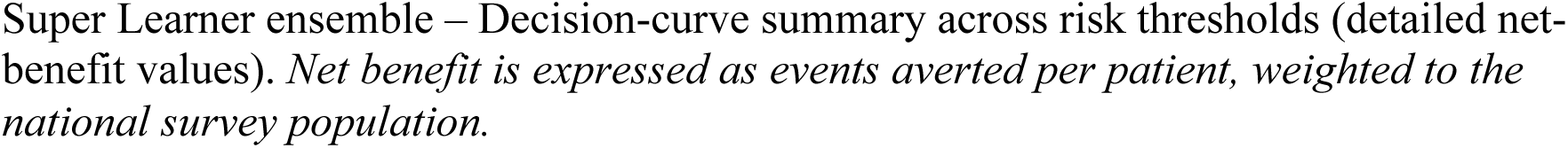
Super Learner ensemble: Decision-curve summary. Super Learner ensemble – Decision-curve summary across risk thresholds (detailed net-benefit values). *Net benefit is expressed as events averted per patient, weighted to the national survey population*.

## Supplementary material

**Figure S6.1.**
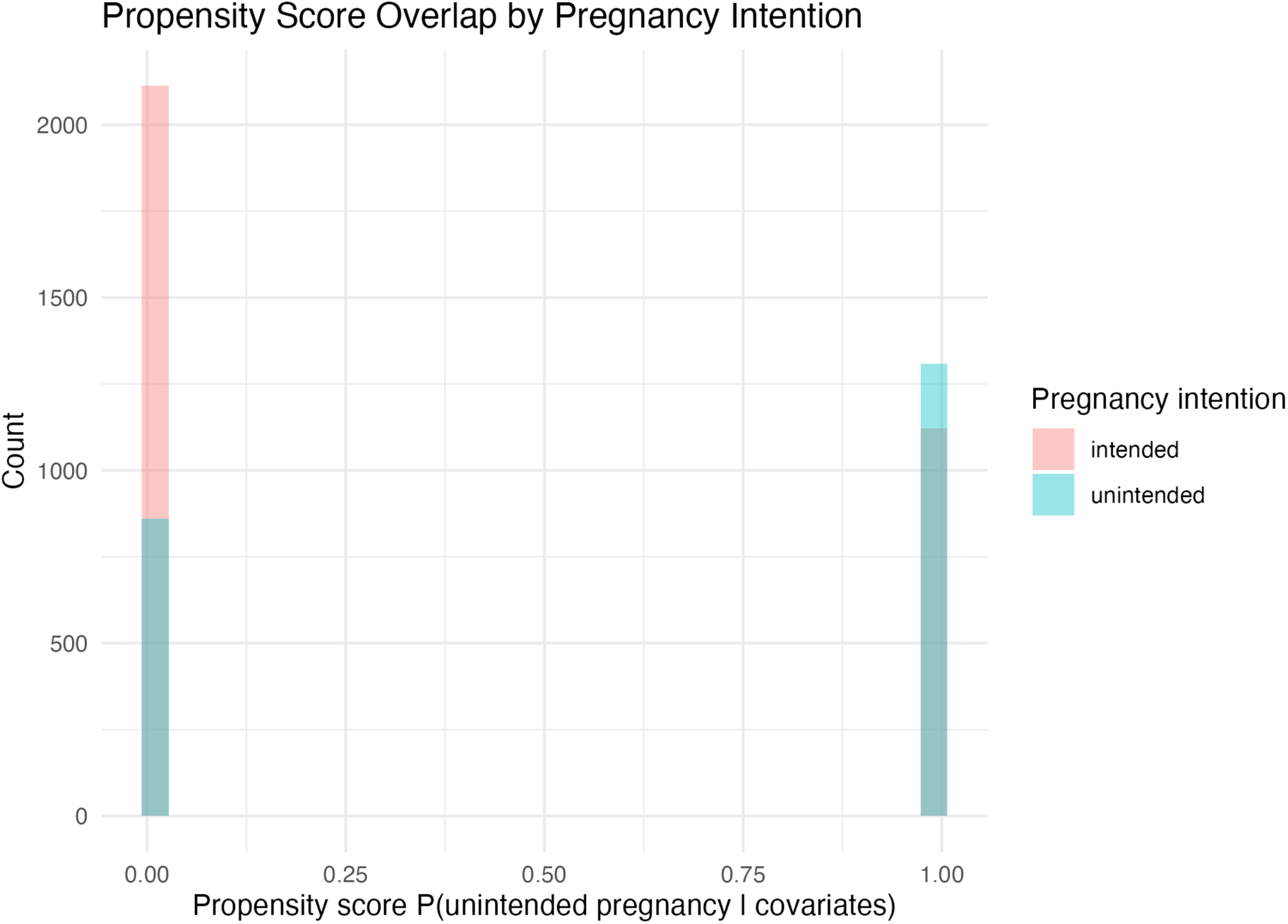
Propensity score overlap histogram by pregnancy intention. (current file: figure6a_ps_hist_overlap.png) *Histogram of estimated propensity scores for unintended pregnancy, stratified by pregnancy intention. Bars show the distribution of scores, highlighting bimodality and limited overlap across groups*.

**Figure S6.2.**
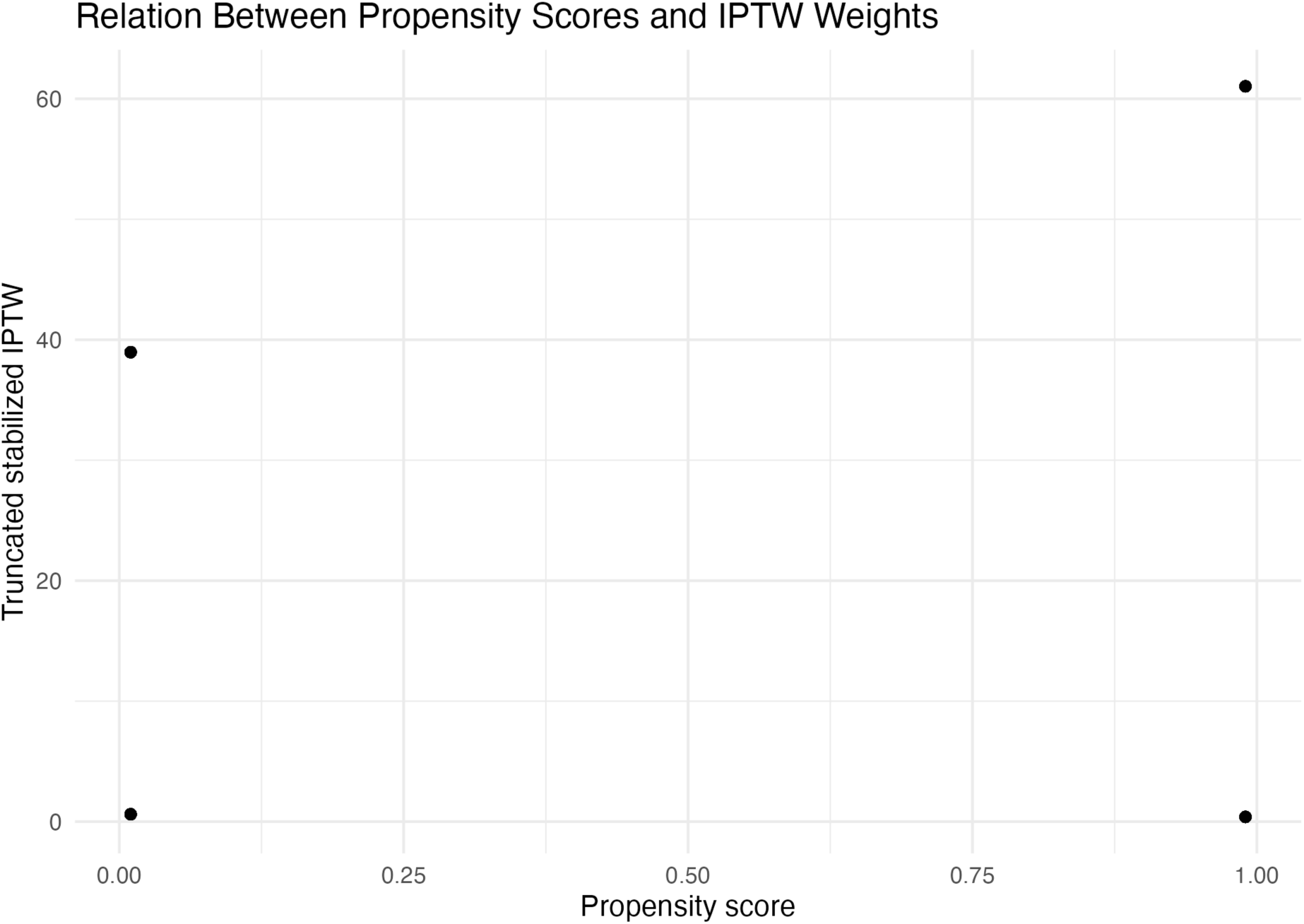
Relation between propensity scores and truncated stabilized IPTW weights. *Scatter plot of estimated propensity scores versus truncated stabilized inverse probability weights. Points demonstrate how extreme propensity scores generate very large weights and how truncation constrains these to improve numerical stability*.

**Table S6.1.**
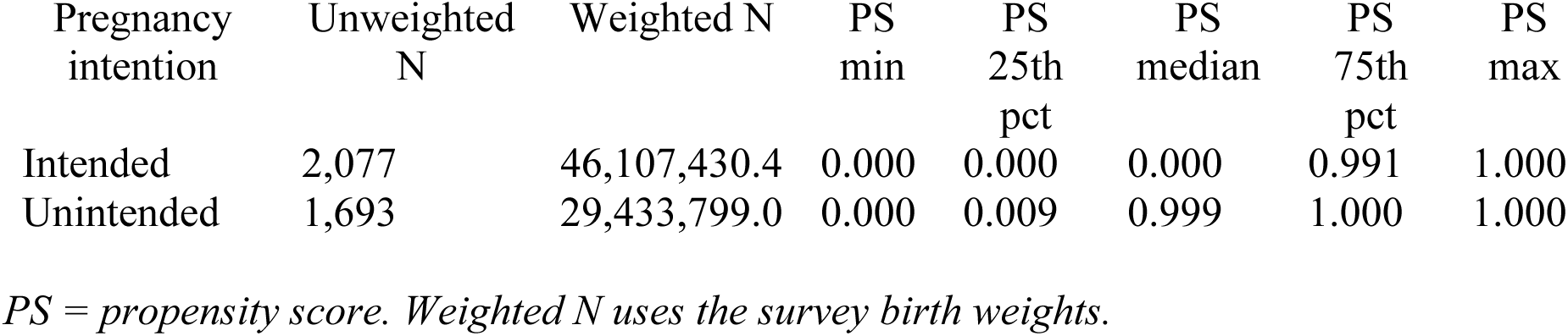
Propensity score summary statistics by pregnancy intention.

**Table S6.2.** E-values for unmeasured confounding.

| Quantity | OR used (treated as RR) | E-value |
| --- | --- | --- |
| Point estimate | 0.701 | 2.207 |
| Confidence interval bound* | 0.058 | — |

E-values for unmeasured confounding. Sensitivity of the doubly robust estimate to a single unmeasured confounder. \*Lower 95% confidence limit; E-value is not defined because the interval includes strong protective effects.

